# Monitoring Red Blood Cell Chimerism with Flow Cytometry

**DOI:** 10.64898/2025.12.11.25341805

**Authors:** ChonHou Ip, Fangming Xiu, Donna Wall

## Abstract

Monitoring the change of cell populations within patients after hematopoietic stem cell transplant (HSCT) is crucial for determining the success of treatment. With current studies largely focused on determining mixed chimerism of nucleated cells, mixed chimerism for non-nucleated red blood cell (RBC) populations was rarely studied. In this study, based on the differences between donor and recipient ABO blood group surface markers and using commercially available mouse monoclonal antibodies (mAbs) for anti-A_1_ and anti-glycophorin A (GlyA), a flow cytometry (FCM) based assay was tested to monitor RBC chimerism for post-HSCT patients with combinations A_1_/O, A_1_B/O, A_1_/B, and A_1_B/B blood group difference. Titration curves of mixed blood type combinations with 10% margins were made. These titration curves had a consistent R^2^ value > 0.99 and a standard deviation (SD) value < 5%. This suggests that the proposed assay was valid, precise, and reliable. Furthermore, a patient sample with recipient A and donor O mixed blood type was tested and showed that the proposed assay was accurate. Since anti-A_2_ and anti-B mAbs were not tested, next steps would be testing these antibodies such that the assay can monitor post-HSCT patients with combinations A_2_/O, A_2_B/O, A_2_/B, A_2_B/B, AB/B, and B/O blood type difference.

## Introduction

RBCs express ABO antigens which define the ABO blood group it belongs to. For a person identified as blood group A, this means that RBCs in the person presents the A antigen. Similarly, a person identified as blood group B will have RBCs with the B antigen. For a person identified as blood group O, RBCs will have no unique antigen. Lastly, for a person identified as blood group AB, RBCs will present both A and B antigen. Across all blood groups, a common surface antigen expressed is glycophorin A (GlyA).^1^ Furthermore, it was commonly found in the serum of individuals identified as blood group A to also have anti-B. For individuals identified as blood group B, they also have anti-A. For individuals identified as blood group O, they have both anti-A and anti-B. For individuals identified as blood group AB, no traces of anti-A and anti-B found.^2^

Under normal circumstances, RBCs within a single human individual will only express one of the four ABO blood groups, however mixed chimerism between blood groups can occur post-HSCT where blood groups are frequently mismatched between donor and recipient. Patients with Thalassemia and Sickle Cell anemia are frequently found to have stable mixed chimerism post-HSCT.^3,4,5^ Stable mixed chimerism is a state where the recipient lymphohematopoietic system consist a mixture of both the host and donor HSCs, where both systems do not reject each other.^6,7^ The ability to monitor the balance between host and donor red cell populations not only become crucial to determine the success of transplant, but may also provide insights for conducting HSCT and solid organ transplant (SOT) in parallel to decrease risk of graft rejection.^8^ Current studies in the field largely focused on determining mixed chimerism of nucleated cells.^8,9,10,11^ Mixed chimerism of non-nucleated RBC populations was rarely studied. Blandhard *et al.* in 1995 presented that FCM can monitor RBC chimerism using mouse mAbs, however they did not conduct a detailed analysis on how precise and accurate FCM can be when used to determine RBCs mixed chimerism.^12^ The current experiment aims to create titration curves of RBC mixed chimerism with 10% margins to determine the precision and accuracy of our FCM based assay using commercially available ABO blood group related mAbs. We tested a patient sample from a child with known mixed donor chimerism.

A challenge in the development of the assay is that there are few antibodies available and they are mostly IgM subtypes - leading to agglutination reactions. Since FCM only detects single cells and agglutination can occur within blood samples, fixation of RBCs was required. Based on unpublished data done by Leung in Wall’s lab, 0.1% of glutaraldehyde (GA) is sufficient to prevent agglutination of RBCs.^13^ In addition, routine tests in Wall’s Lab discovered that binding affinity of anti-A varies greatly across blood A and AB. This can be due to the sub-groups of A_1_ and A_2_.^14,15^ To determine the specificity of anti-A used in this experiment, preliminary agglutination test using anti-A_1_ lectin was done. Results showed that anti-A used in this experiment was specific to the blood groups A_1_ and A_1_B.

## Material and Methods

### Preparing red blood cell samples

This study was approved by SickKids Research Ethics Board (REB). Healthy donor blood samples for each blood group and patient blood sample (recipient A, donor O) were collected from the Toronto SickKids Hospital. To avoid excessive hemolysis and blood agglutination, fresh blood samples stored under 4 ^0^C was renewed every two weeks. For blood samples that contains group A or AB, agglutination test was done with anti-A_1_ lectin and only A_1_ positive blood samples were used for the experiment.

In preparation for immunostaining, 10 ηl of blood sample was diluted 100x using 0,1% saline. Using the Z2 Coulter Particle counter and cell analyzer (Beckman Coulter, US), the concentration of blood sample was determined. Artificially mixed chimerism blood samples (A_1_/O, A_1_B/O, A_1_/B, and A_1_B/B) with 10% increments were made by mixing relative percentages of the two blood groups of interest. This creates RBC mixed chimerism samples with a total of 10 million cells. To prevent agglutination of cells, the sample prepared was fixated with 0.1% aqueous GA (Thermo scientific, Cat: A17876.AE, Lot: 10222932) for 10 minutes.

The sample was then washed with 2 mL of saline and centrifuged at 600g for 5 minutes twice. It is important to note that throughout the experiment, sample with RBCs was placed under vortex for at least 5 seconds before each transfer.

### Antibodies

Antibodies were titrated before being used. For BV421 Mouse Anti-Human CD235a (anti-GlyA) (BD Horizon™, Cat: 562938, Lot#: 1334017), RBC and isolated PBMC were used to determine the titrated volume of 2 ηl. For Alexa Fluor^®^ 647 Mouse Anti-Human Blood Group A (anti-A) (BD Pharmingen™, Cat: 565384, Lot#: 2342371), RBC of blood group A_1_ and blood group B were used to determine the titrated volume of 5 ηl.

### Flow cytometry

For each sample, 1 million cells resuspended in 50 ηl of saline was used for staining. An unstained sample was used as a control. After adding the corresponding titrated volumes of both anti-GlyA and anti-A, the sample was incubated for 15 minutes at room temperature or 30 minutes at 4 ^0^C. FCM (LSR II GC) was used to screen all samples. Data collected were analyzed using the software FlowJo (V10) where the 10 thousand events recorded was gated on RBCs. With doublet discrimination performed, events were further gated only on anti-GlyA positive cells, and lastly represented on a graph with anti-GlyA against anti-A. The gates for positive and negative anti Gly-A and anti-A were determined based on the unstained sample. The relative percentages shown on the double positive and single positive gates were recorded and analyzed.

Each combination was tested 5 times to obtain the mean and a SD value at each 10% margin. The patient sample was only tested 3 times. Repeats of the experiment began on determining the concentration of the sample. This ensures that SD values reflects the sum of all instrumental error that exists throughout the experiment.

## Results

### Artificially mixed RBC chimerism samples

Based on Figure 1 and Table 1, it was observed that combination A_1_/O gave average values that forms a titrated curve with an R^2^ value of 0.9982 and 0.9983. The average SD was 2.5% for blood group A_1_ and 2.7% for blood group O. Respective FCM plots can be found in Supplementary Figures 1.1, 1.2, 1.3, 1.4, and 1.5. On Figure 2 and Table 2, it was observed that combination A_1_B/O gave average values that forms a titrated curve with an R^2^ value of 0.9996. The average SD for both blood groups A_1_B and O was 1.5%. Respective FCM plots can be found in Supplementary Figures 2.1, 2.2, 2.3, 2.4, and 2.5. Furthermore, on Figure 3 and Table 3, it was observed that combination A_1_/B gave average values that forms a titrated curve with an R^2^ value of 0.9996. The average SD for both blood groups A_1_ and B was 1.7%. Respective FCM plots can be found in Supplementary Figures 3.1, 3.2, 3.3, 3.4, and 3.5. Lastly, on Figure 4 and Table 4, it was observed that combination A_1_B /B gave average values that forms a titrated curve with an R^2^ value of 0.9983. The average SD was 1.6% for blood group AB and 1.5% for blood group B. Respective FCM plots can be found in Supplementary Figures 4.1, 4.2, 4.3, 4.4, and 4.5.

**Figure 1:**
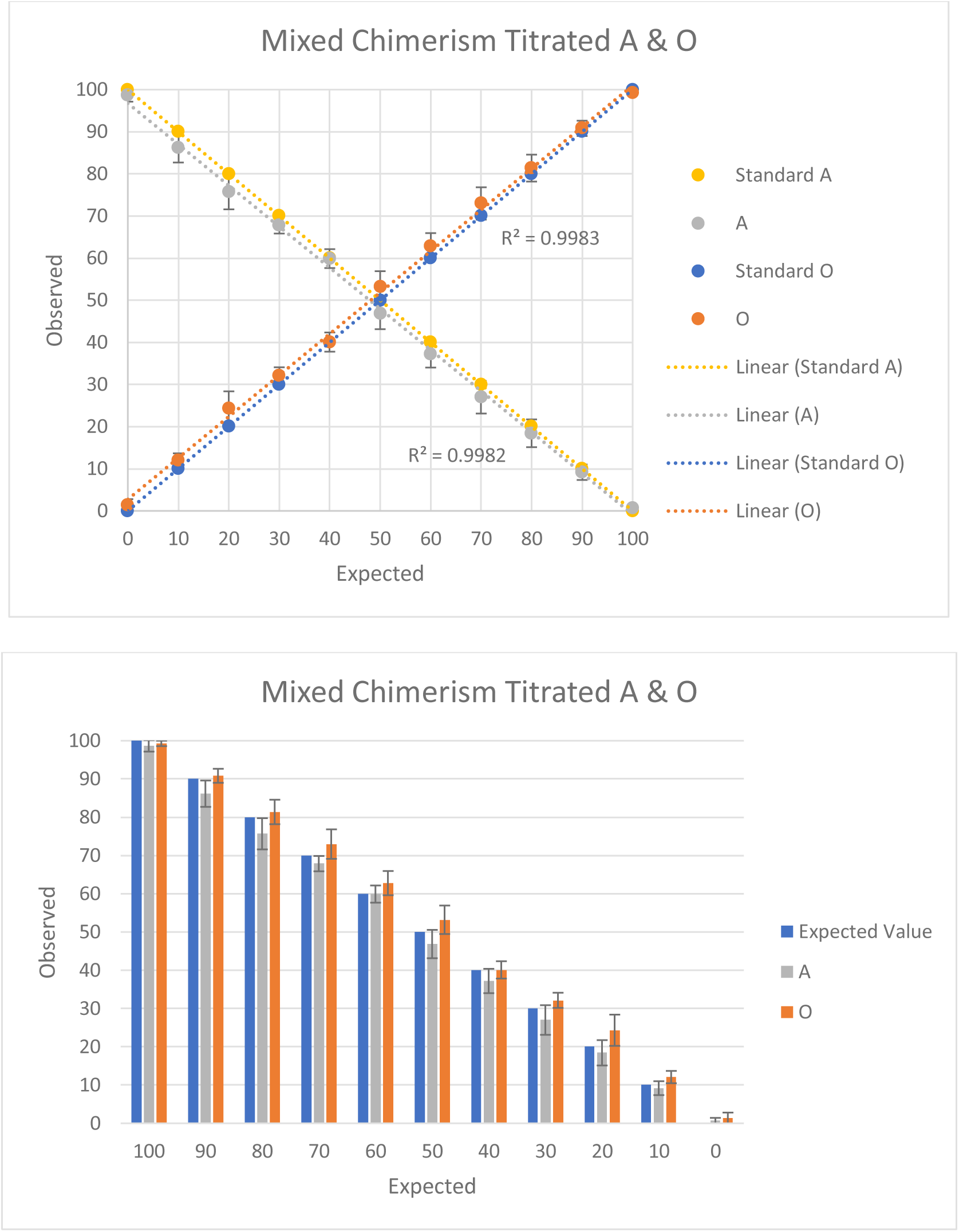
Titration curve with 10% margins for artificially mixed chimerism combination A_1_/O. Mean values of 5 repeated experiments are plotted on a graph along with the calculated standard deviation shown for each data point. The bar graph shows the same data but provides an alternative visual presentation of the data.

**Figure 2:**
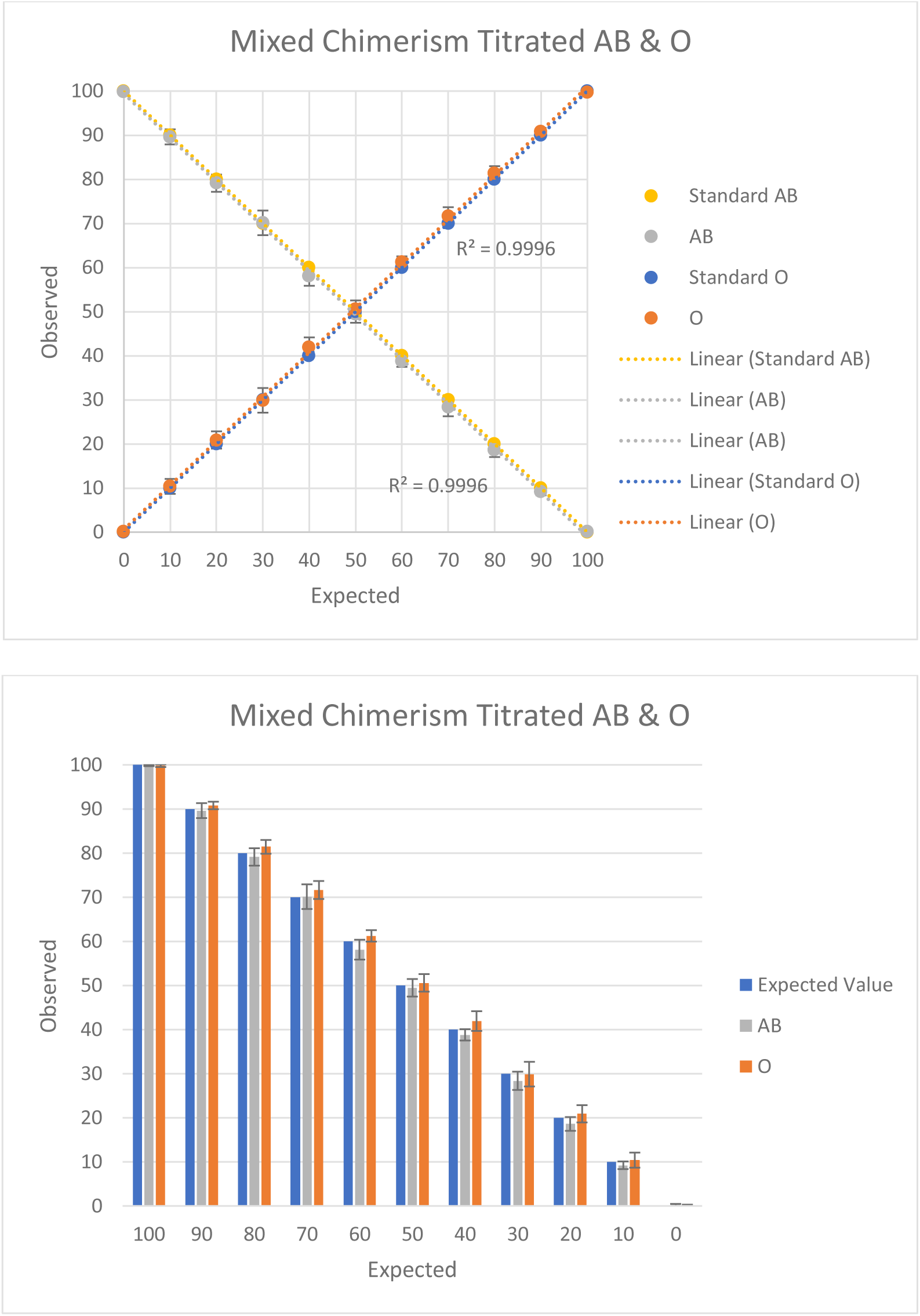
Titration curve with 10% margins for artificially mixed chimerism combination A_1_B/O. Mean values of 5 repeated experiments are plotted on a graph along with the calculated standard deviation shown for each data point. The bar graph shows the same data but provides an alternative visual presentation of the data.

**Figure 3:**
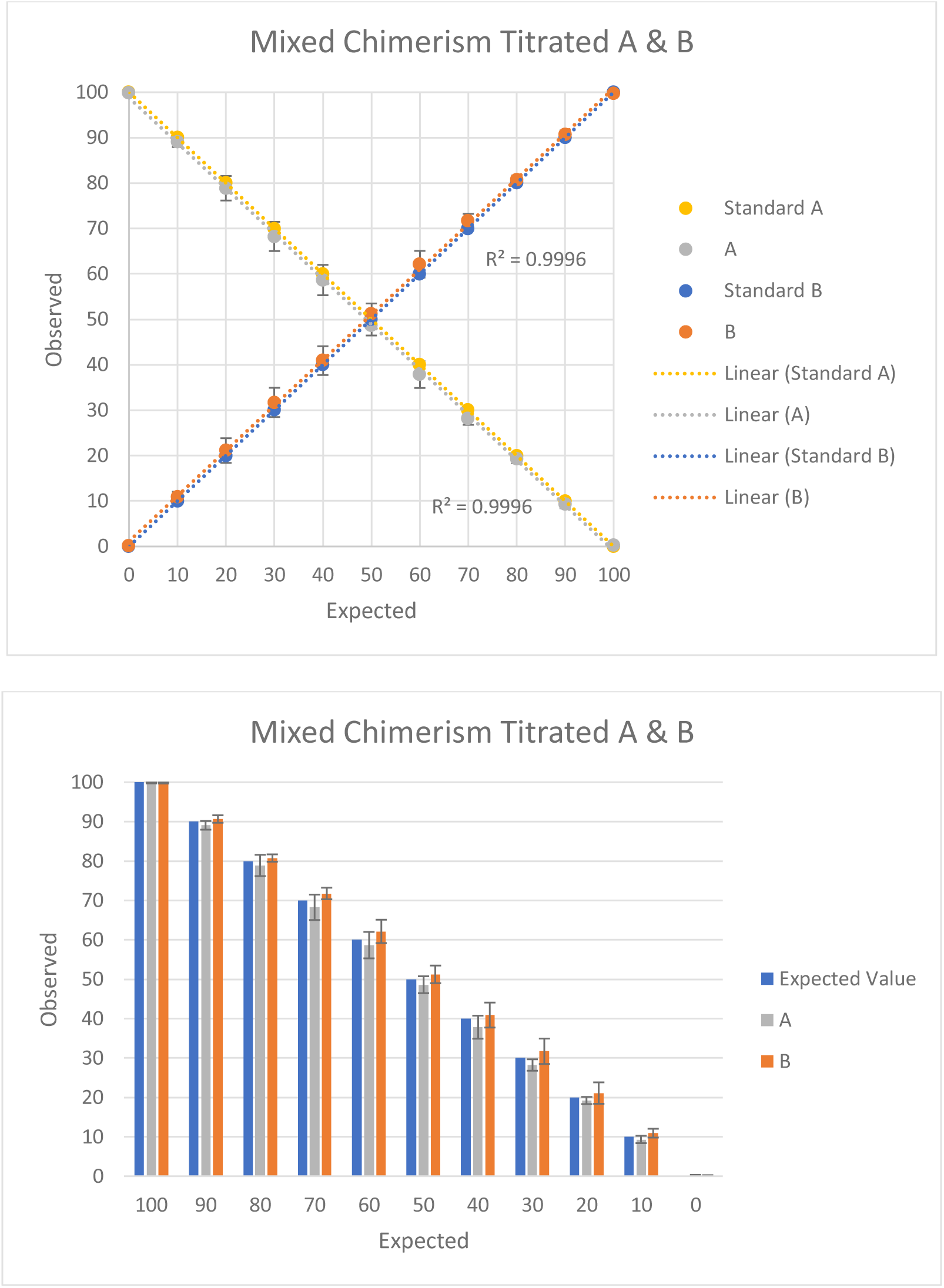
Titration curve with 10% margins for artificially mixed chimerism combination A_1_/B. Mean values of 5 repeated experiments are plotted on a graph along with the calculated standard deviation shown for each data point. The bar graph shows the same data but provides an alternative visual presentation of the data.

**Figure 4:**
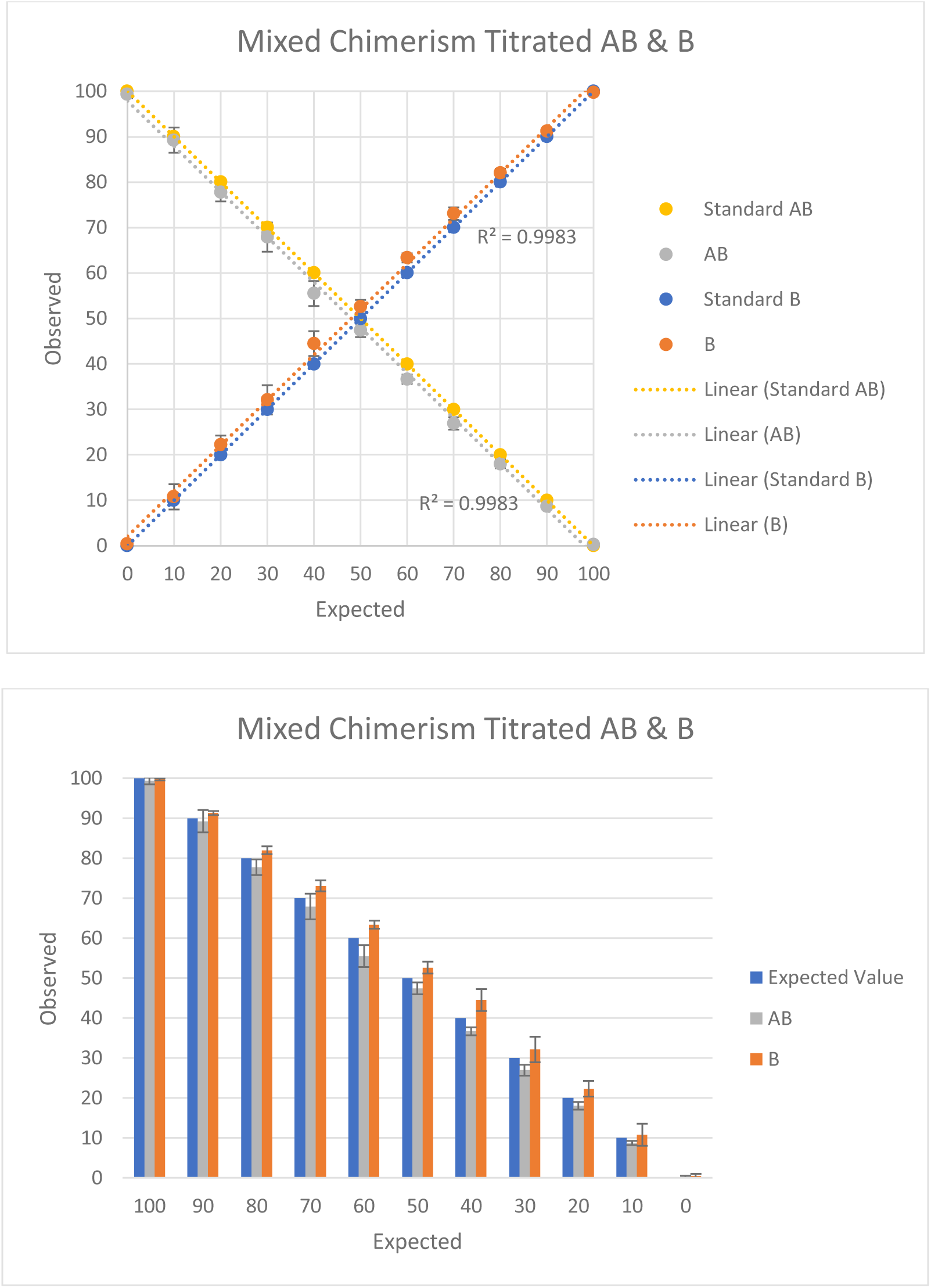
Titration curve with 10% margins for artificially mixed chimerism combination A_1_B/B. Mean values of 5 repeated experiments are plotted on a graph along with the calculated standard deviation shown for each data point. The bar graph shows the same data but provides an alternative visual presentation of the data.

**Table 1:** Analysis on Flow data for mixed chimerism A_1_/O. Table shows extracted values from FCM results illustrated in Supplementary Figure 1.1, 1.2, 1.3, 1.4 and 1.5. Based on the collected data, the mean and the standard deviation values were calculated for each 10% increment and plotted as a line and bar graph presented on Figure 1.

| Titration A & O |  |  |  |  |  |  |  |  |  |  |  |
| --- | --- | --- | --- | --- | --- | --- | --- | --- | --- | --- | --- |
| Expected value | 100 | 90 | 80 | 70 | 60 | 50 | 40 | 30 | 20 | 10 | 0 |
| Flow data | A |  |  |  |  |  |  |  |  |  |  |
| Date performed |  |  |  |  |  |  |  |  |  |  |  |
| 2023-01-23 | 99.7 | 86.8 | 76.6 | 65.1 | 62.2 | 43.6 | 36.6 | 23.7 | 16.7 | 8.69 | 0.43 |
| 2023-02-02 | 97.0 | 80.5 | 68.6 | 69.9 | 61.1 | 52.3 | 40.6 | 32.5 | 23.2 | 11.3 | 0.81 |
| 2023-02-09 | 99.8 | 89.7 | 78.3 | 69.7 | 61.2 | 48.2 | 39.0 | 28.9 | 20.0 | 10.7 | 0.2 |
| 2023-02-23 | 99.7 | 87.6 | 76.3 | 67.8 | 57.2 | 43.3 | 32.2 | 23.1 | 14.6 | 6.78 | 0.2 |
| 2023-03-27 | 97.0 | 86.1 | 78.5 | 66.8 | 57.8 | 46.8 | 37.6 | 26.8 | 17.6 | 8.39 | 1.87 |
| Mean | 98.6 | 86.1 | 75.7 | 67.9 | 59.9 | 46.8 | 37.2 | 27.0 | 18.4 | 9.2 | 0.7 |
| SD | 1.5 | 3.4 | 4.1 | 2.0 | 2.2 | 3.7 | 3.2 | 3.9 | 3.3 | 1.8 | 0.7 |
| Flow data | O |  |  |  |  |  |  |  |  |  |  |
| Date performed |  |  |  |  |  |  |  |  |  |  |  |
| 2023-01-23 | 99.5 | 91.3 | 82.3 | 76.2 | 63.3 | 56.5 | 37.7 | 34.8 | 23.3 | 13.2 | 0.26 |
| 2023-02-02 | 99.2 | 88.7 | 76.8 | 67.5 | 59.4 | 47.7 | 38.9 | 30.1 | 31.4 | 10.5 | 2.84 |
| 2023-02-09 | 99.8 | 89.3 | 80 | 71.1 | 61 | 51.8 | 38.8 | 30.3 | 21.7 | 10.3 | 0.2 |
| 2023-02-23 | 99.8 | 93.2 | 85.4 | 76.9 | 67.8 | 56.7 | 42.8 | 32.2 | 23.7 | 12.4 | 0.27 |
| 2023-03-27 | 98.1 | 91.6 | 82.4 | 73.2 | 62.4 | 53.2 | 42.2 | 33.2 | 21.5 | 13.9 | 3.0 |
| Mean | 99.3 | 90.8 | 81.4 | 73.0 | 62.8 | 53.2 | 40.1 | 32.1 | 24.3 | 12.1 | 1.3 |
| SD | 0.7 | 1.8 | 3.2 | 3.9 | 3.2 | 3.7 | 2.3 | 2.0 | 4.1 | 1.6 | 1.5 |

**Table 2:**
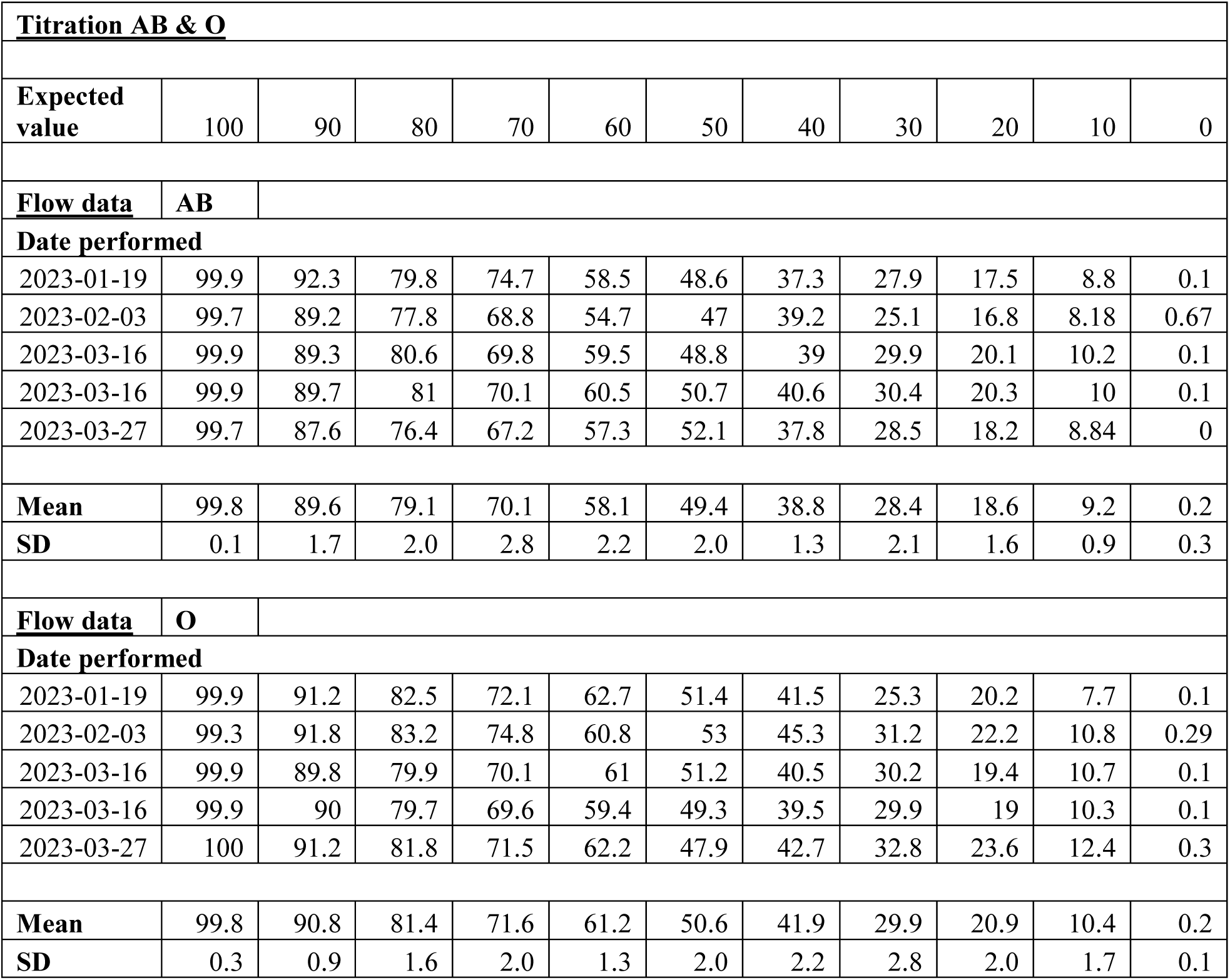
Analysis on Flow data for mixed chimerism A_1_B/O. Table shows extracted values from FCM results illustrated in Supplementary Figure 2.1, 2.2, 2.3, 2.4 and 2.5. Based on the collected data, the mean and the standard deviation values were calculated for each 10% increment and plotted as a line and bar graph presented on Figure 2.

| Titration AB & O |  |  |  |  |  |  |  |  |  |  |  |
| --- | --- | --- | --- | --- | --- | --- | --- | --- | --- | --- | --- |
| Expected value | 100 | 90 | 80 | 70 | 60 | 50 | 40 | 30 | 20 | 10 | 0 |
| Flow data | AB |  |  |  |  |  |  |  |  |  |  |
| Date performed |  |  |  |  |  |  |  |  |  |  |  |
| 2023-01-19 | 99.9 | 92.3 | 79.8 | 74.7 | 58.5 | 48.6 | 37.3 | 27.9 | 17.5 | 8.8 | 0.1 |
| 2023-02-03 | 99.7 | 89.2 | 77.8 | 68.8 | 54.7 | 47 | 39.2 | 25.1 | 16.8 | 8.18 | 0.67 |
| 2023-03-16 | 99.9 | 89.3 | 80.6 | 69.8 | 59.5 | 48.8 | 39 | 29.9 | 20.1 | 10.2 | 0.1 |
| 2023-03-16 | 99.9 | 89.7 | 81 | 70.1 | 60.5 | 50.7 | 40.6 | 30.4 | 20.3 | 10 | 0.1 |
| 2023-03-27 | 99.7 | 87.6 | 76.4 | 67.2 | 57.3 | 52.1 | 37.8 | 28.5 | 18.2 | 8.84 | 0 |
| Mean | 99.8 | 89.6 | 79.1 | 70.1 | 58.1 | 49.4 | 38.8 | 28.4 | 18.6 | 9.2 | 0.2 |
| SD | 0.1 | 1.7 | 2.0 | 2.8 | 2.2 | 2.0 | 1.3 | 2.1 | 1.6 | 0.9 | 0.3 |
| Flow data | O |  |  |  |  |  |  |  |  |  |  |
| Date performed |  |  |  |  |  |  |  |  |  |  |  |
| 2023-01-19 | 99.9 | 91.2 | 82.5 | 72.1 | 62.7 | 51.4 | 41.5 | 25.3 | 20.2 | 7.7 | 0.1 |
| 2023-02-03 | 99.3 | 91.8 | 83.2 | 74.8 | 60.8 | 53 | 45.3 | 31.2 | 22.2 | 10.8 | 0.29 |
| 2023-03-16 | 99.9 | 89.8 | 79.9 | 70.1 | 61 | 51.2 | 40.5 | 30.2 | 19.4 | 10.7 | 0.1 |
| 2023-03-16 | 99.9 | 90 | 79.7 | 69.6 | 59.4 | 49.3 | 39.5 | 29.9 | 19 | 10.3 | 0.1 |
| 2023-03-27 | 100 | 91.2 | 81.8 | 71.5 | 62.2 | 47.9 | 42.7 | 32.8 | 23.6 | 12.4 | 0.3 |
| Mean | 99.8 | 90.8 | 81.4 | 71.6 | 61.2 | 50.6 | 41.9 | 29.9 | 20.9 | 10.4 | 0.2 |
| SD | 0.3 | 0.9 | 1.6 | 2.0 | 1.3 | 2.0 | 2.2 | 2.8 | 2.0 | 1.7 | 0.1 |

**Table 3:** Analysis on Flow data for mixed chimerism A_1_/B. Table shows extracted values from FCM results illustrated in Supplementary Figure 3.1, 3.2, 3.3, 3.4 and 3.5. Based on the collected data, the mean and the standard deviation values were calculated for each 10% increment and plotted as a line and bar graph presented on Figure 3.

| Titration A & B |  |  |  |  |  |  |  |  |  |  |  |
| --- | --- | --- | --- | --- | --- | --- | --- | --- | --- | --- | --- |
| Expected value | 100 | 90 | 80 | 70 | 60 | 50 | 40 | 30 | 20 | 10 | 0 |
| Flow data | A |  |  |  |  |  |  |  |  |  |  |
| Date performed |  |  |  |  |  |  |  |  |  |  |  |
| 2023-01-23 | 99.9 | 89.3 | 79.4 | 72 | 59.3 | 48.8 | 41 | 30 | 19.7 | 9.6 | 0.4 |
| 2023-01-26 | 99.7 | 89.3 | 78.9 | 70.1 | 56.7 | 49.3 | 39.4 | 29.6 | 20.1 | 10.6 | 0.29 |
| 2023-03-01 | 99.7 | 88.5 | 78.9 | 67.8 | 55.5 | 46.2 | 36.2 | 27.6 | 17.7 | 8.4 | 0.2 |
| 2023-03-02 | 99.9 | 87.6 | 82.4 | 63.4 | 64.1 | 51.7 | 33.6 | 27.3 | 19.4 | 8.44 | 0.1 |
| 2023-03-06 | 99.9 | 90.6 | 74.8 | 68 | 57.7 | 47 | 39 | 26.7 | 19.2 | 9.6 | 0.1 |
| Mean | 99.8 | 89.1 | 78.9 | 68.3 | 58.7 | 48.6 | 37.8 | 28.2 | 19.2 | 9.3 | 0.2 |
| SD | 0.1 | 1.1 | 2.7 | 3.2 | 3.3 | 2.1 | 2.9 | 1.5 | 0.9 | 0.9 | 0.1 |
| Flow data | B |  |  |  |  |  |  |  |  |  |  |
| Date performed |  |  |  |  |  |  |  |  |  |  |  |
| 2023-01-23 | 99.6 | 90.4 | 80.3 | 70 | 59 | 51.2 | 40.7 | 28 | 20.6 | 10.7 | 0.1 |
| 2023-01-26 | 99.7 | 89.4 | 79.9 | 70.4 | 60.5 | 49.9 | 41.2 | 29.8 | 21.1 | 10.7 | 0.25 |
| 2023-03-01 | 99.8 | 91.6 | 82.3 | 72.4 | 63.8 | 53.8 | 44.5 | 32.2 | 21.1 | 11.5 | 0.3 |
| 2023-03-02 | 99.9 | 91.6 | 80.6 | 72.7 | 66.4 | 48.3 | 35.9 | 36.6 | 17.6 | 12.4 | 0.1 |
| 2023-03-06 | 99.9 | 90.3 | 80.7 | 73.3 | 61 | 53 | 42.3 | 32 | 25.2 | 9.4 | 0.1 |
| Mean | 99.8 | 90.7 | 80.8 | 71.8 | 62.1 | 51.2 | 40.9 | 31.7 | 21.1 | 10.9 | 0.2 |
| SD | 0.1 | 0.9 | 0.9 | 1.5 | 2.9 | 2.2 | 3.2 | 3.2 | 2.7 | 1.1 | 0.1 |

**Table 4:** Analysis on Flow data for mixed chimerism A_1_B/B. Table shows extracted values from FCM results illustrated in Supplementary Figure 4.1, 4.2, 4.3, 4.4 and 4.5. Based on the collected data, the mean and the standard deviation values were calculated for each 10% increment and plotted as a line and bar graph presented on Figure 4.

| Titration AB & B |  |  |  |  |  |  |  |  |  |  |  |
| --- | --- | --- | --- | --- | --- | --- | --- | --- | --- | --- | --- |
| Expected value | 100 | 90 | 80 | 70 | 60 | 50 | 40 | 30 | 20 | 10 | 0 |
| Flow data | AB |  |  |  |  |  |  |  |  |  |  |
| Date performed |  |  |  |  |  |  |  |  |  |  |  |
| 2023-01-20 | 97.8 | 88.2 | 78.6 | 69.4 | 54.7 | 45.4 | 35.1 | 28.1 | 19.3 | 8.45 | 0.66 |
| 2023-03-15 | 99.7 | 87.3 | 78.8 | 72.8 | 52.4 | 49.3 | 37.2 | 27.2 | 17.8 | 8.97 | 0.32 |
| 2023-03-15 | 99.7 | 89 | 79 | 65.1 | 58.8 | 47.9 | 37.3 | 27.1 | 18.2 | 9.5 | 0.1 |
| 2023-03-15 | 99.8 | 94.1 | 74.3 | 65.5 | 53.7 | 47.9 | 36.2 | 24.6 | 18.2 | 8.3 | 0.1 |
| 2023-03-16 | 99.7 | 87.6 | 77.9 | 66.7 | 57.9 | 46.5 | 37.5 | 27.6 | 16.6 | 8.3 | 0.1 |
| Mean | 99.3 | 89.2 | 77.7 | 67.9 | 55.5 | 47.4 | 36.7 | 26.9 | 18.0 | 8.7 | 0.3 |
| SD | 0.9 | 2.8 | 2.0 | 3.2 | 2.7 | 1.5 | 1.0 | 1.4 | 1.0 | 0.5 | 0.2 |
| Flow data | B |  |  |  |  |  |  |  |  |  |  |
| Date performed |  |  |  |  |  |  |  |  |  |  |  |
| 2023-01-20 | 99.3 | 91.5 | 80.7 | 71.8 | 64.9 | 54.6 | 45.2 | 30.6 | 21.4 | 11.8 | 1.4 |
| 2023-03-15 | 99.7 | 91 | 82.2 | 72.8 | 62.8 | 50.7 | 47.6 | 27.2 | 21.2 | 12.7 | 0.14 |
| 2023-03-15 | 99.9 | 90.5 | 81.8 | 72.9 | 62.7 | 52.1 | 41.2 | 34.9 | 21 | 11 | 0.26 |
| 2023-03-15 | 99.9 | 91.7 | 81.8 | 75.4 | 63.8 | 52.1 | 46.3 | 34.5 | 25.7 | 5.93 | 0.2 |
| 2023-03-16 | 99.9 | 91.7 | 83.4 | 72.4 | 62.5 | 53.5 | 42.1 | 33.3 | 22.1 | 12.4 | 0.2 |
| Mean | 99.7 | 91.3 | 82.0 | 73.1 | 63.3 | 52.6 | 44.5 | 32.1 | 22.3 | 10.8 | 0.4 |
| SD | 0.3 | 0.5 | 1.0 | 1.4 | 1.0 | 1.5 | 2.7 | 3.2 | 2.0 | 2.8 | 0.5 |

### Patient blood sample

A patient who had stable mixed chimerism 18 months post unrelated donor transplant for treatment of an immunodeficiency syndrome was tested for chimerism of RBCs. Based on Figure 5 and Table 5, the patient sample that consist a mixture of donor blood group O and recipient blood group A was shown to be 78.1% group O blood and 21.9% group A blood, both with a SD of 0.6%. Respective FCM plots can be found in Supplementary Figure 5.1.

**Figure 5:**
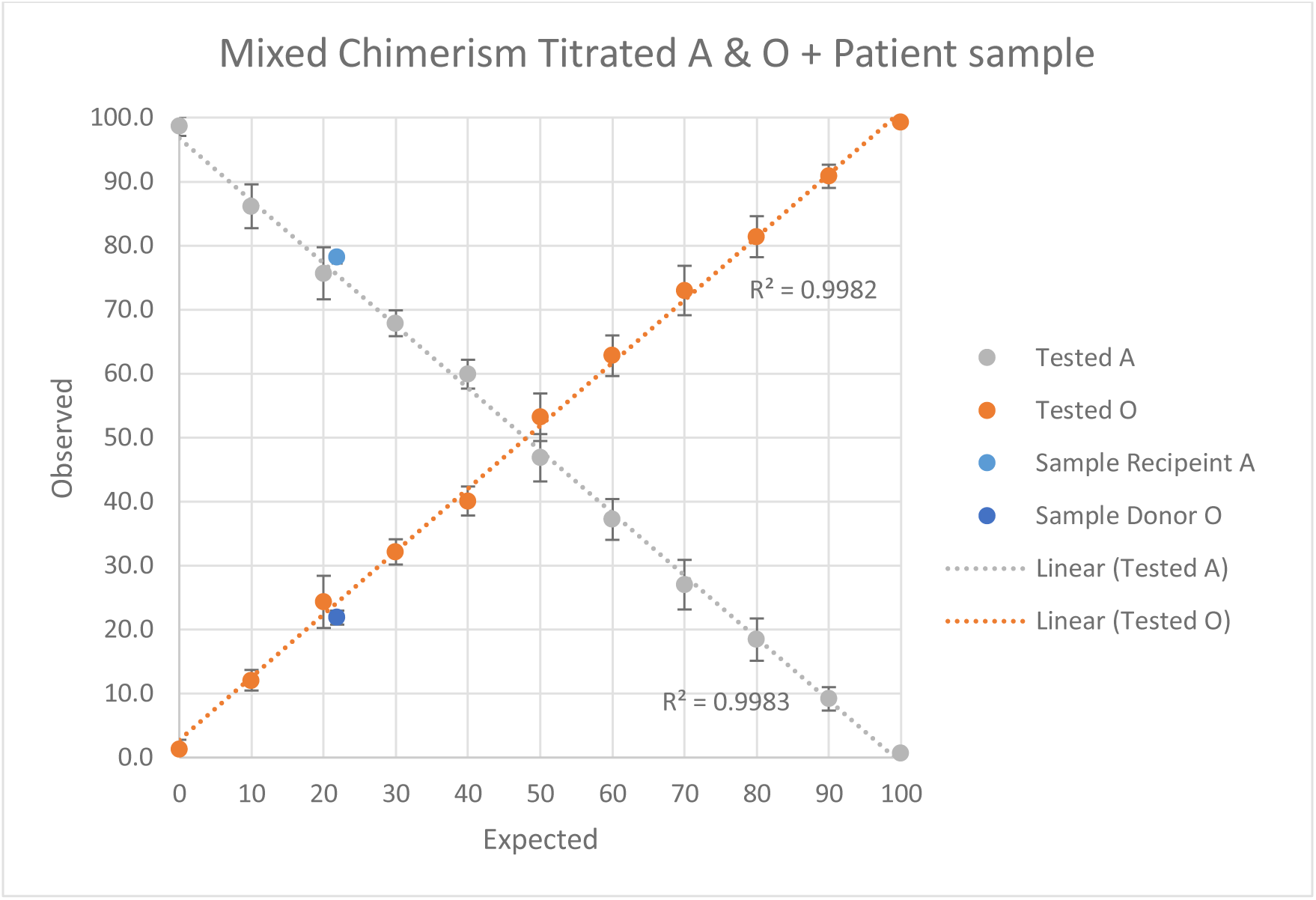
Patient sample plotted on the titration curve for artificially mixed chimerism combination A_1_/O. Mean values of 5 repeated experiments are plotted on a graph along with the calculated SD shown for each data point. Patient sample with mixed chimerism of recipient A and donor O are also potted on the same graph showing their respective locations.

**Table 5:** Analysis on Patient sample with mixed chimerism Recipient A_1_ and Donor O. Table shows extracted values from flow cytometry results illustrated in Supplementary Figure 5.1. Based on the collected data from the triplet experiment, the mean and the SD values were calculated. Data points in combination with Figure 1 were plotted on Figure 5.

| **Patient sample 1** |  |  |  |  |  |
| --- | --- | --- | --- | --- | --- |
| **Blood group** | **Data 1** | **Data 2** | **Data 3** | **Mean** | **SD** |
| Recipient A<sub>1</sub> | 77.6 | 78 | 78.8 | 78.1 | 0.6 |
| Donor O | 22.4 | 22 | 21.2 | 21.9 | 0.6 |

## Discussion

### Proposed RBC mixed chimerism flow cytometry assay

With a consistent R^2^ value greater than 0.99 for all titration curves and a consistent SD value less than 5%, this suggests that for artificial mixed chimerism combinations A_1_/O, A_1_B/O, A_1_/B, and A_1_B/B, the proposed FCM based assay using commercially available anti-A_1_ and anti-GlyA was able to monitor RBC chimerism with precision. Moreover, with consistent results shown across repeated tests on mixing studies, this also showed that the proposed assay was reliable. Moreover, with the unstained samples tested negative for anti-GlyA and anti-A, detection of 100% blood group A_1_ and A_1_B using anti-A_1_ was approximately 99%, the detection of 100% blood group O and B was also approximately 99%, and the above results stayed consistent across all repeated tests, this suggests that the design of the experiment was valid where mAbs used detects the intended blood groups within the mixed chimerism sample.

Furthermore, with the patient blood sample located within the SD range of the titration curve for mixed chimerism A_1_/O, this suggests that the FCM based proposed assay to monitor RBC chimerism was accurate for detecting blood group combination A_1_/O.

### Comparison with other nucleated cells

On the same day when RBC chimerism of patient sample was tested, a chimerism test for nucleated cells in the patient sample was done. Unpublished data showed that chimerism of myeloid (CD33) and lymphoid (CD3) was 14% and 51% donor respectively. Based on these data, although it can be observed that RBC chimerism more closely reflected on myeloid cells, it also suggests that chimerism is not consistent among different cell types.

### Limitations and Improvements

Although the proposed FCM assay to monitor RBC chimerism was shown to be precise, reliable, valid and accurate, with only one patient sample tested, the degree of accuracy for the proposed FCM assay remain to be determined. Increasing the number of patient samples tested may further support the accuracy of the proposed FCM assay to monitor RBC chimerism.

Moreover, commercial mAbs for anti-A_2_ and anti-B was not tested in this experiment which limits the current proposed FCM based assay to be applied for the remaining blood group combinations A_2_/O, A_2_B/O, A_2_/B, A_2_B/B, AB/B, and B/O. Therefore, a step further to broaden the coverage of the FCM based chimerism assay would be to repeat the experiment using commercial mAbs for anti-A_2_ and anti-B, test the remaining blood chimerism combinations, and to test the accuracy of the assay using real patient samples.

### Application of assay

With the proposed FCM based assay able to monitor RBC chimerism with precision while providing valid, reliable and accurate results, this assay can help clinicians to monitor RBC mixed chimerism in patients suffering from Thalassemia and Sickle Cell anemia where the survival of donor red blood cells is longer than recipient cells. Knowledge of the degree of chimerism will help with clinical management.

As SOT strategies include a strategy of inducing organ tolerance, low levels of mixed donor recipient hematopoietic chimerism is applied. There are unique issues in SOT when ABO blood group mismatch is involved. With vascular endothelial cells of the donor organ expressing blood group antigens, pre-existing anti-A or anti-B antibodies in the recipient can lead to hyperacute rejection.^16^ In most transplants, it was found that immunological tolerance is closely associated with T regulatory (Treg) cells.^17^ Therefore, the ability to reliably monitor donor and recipient cell populations is significant to provide further insights into the nature of stable mixed chimerism state and to determine whether parallel HSCT and SOT is plausible to minimize risk of graft rejection.

Currently, the clinical lab team in SickKids has ongoing studies that will require a reliable assay to monitor RBC chimerism. The Thal-FabS study directed by Dr. Kuang-Yueh Chiang involves a secondary objective of monitoring cell chimerism post-HSCT. On the other hand, another study directed by Dr. Joerg Krueger involves the objective of investigating how stable mixed chimerism was achieved in ABO mismatched transplant patients. The proposed FCM-based RBC chimerism assay in this study may help the ongoing studies to achieve their respective objectives.

## Data Availability

All data produced in the present study are available upon reasonable request to the authors

## Acknowledgements

The authors would like to thank all members of Dr. Donna Wall’s lab and the University of Toronto Immunology department. The authors also thank previous and current members of project including Raymond Kung, Karin G. Hermans, Ginnian Leung, Ada Tsang, ZhiJuan Luo and Rachel Tse.

## Supplementary Figures

**Supplementary Figure 1.1:**
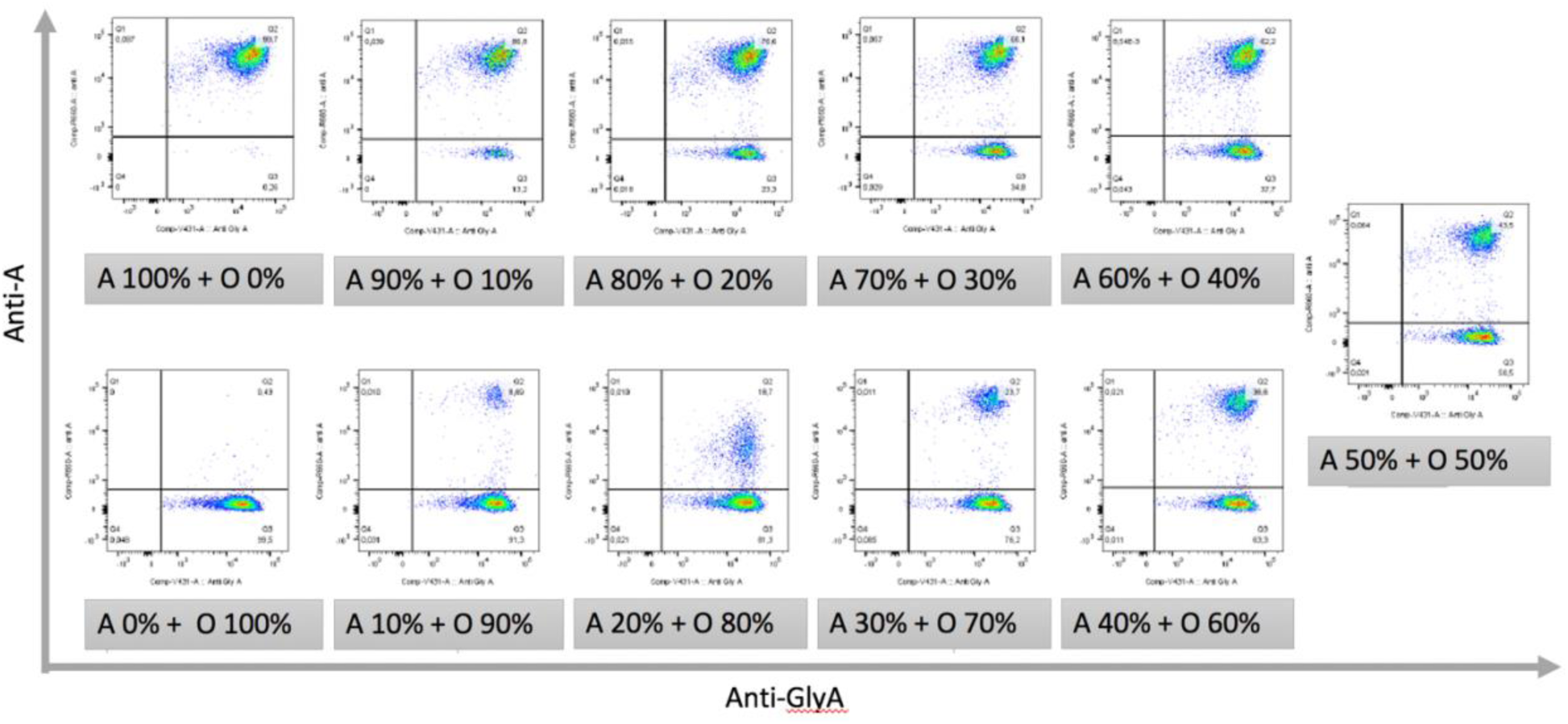
Flow cytometry data for A_1_/O – 2023-01-23. Data obtained through using the proposed FCM-based RBC chimerism assay for artificially mixed combination of blood groups A_1_/O. The experiment was carried out on 2023-01-23.

**Supplementary Figure 1.2:**
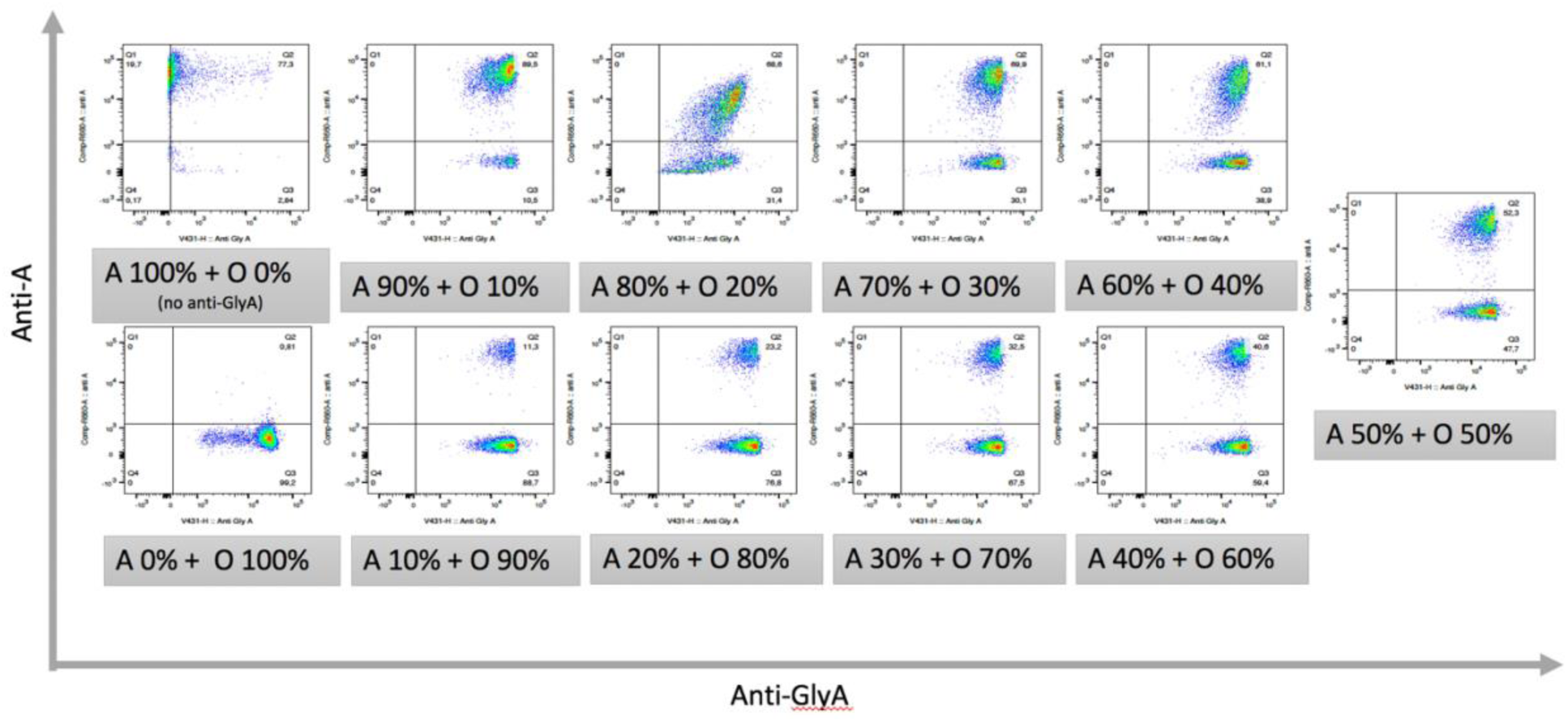
Flow cytometry data for A_1_/O – 2023-02-02. Data obtained through using the proposed FCM-based RBC chimerism assay for artificially mixed combination of blood groups A_1_/O. The experiment was carried out on 2023-02-02. Since combination A100% + O 0% was not stained with anti-GlyA, values obtained are only based on positive anti-A and negative anti-A.

**Supplementary Figure 1.3:**
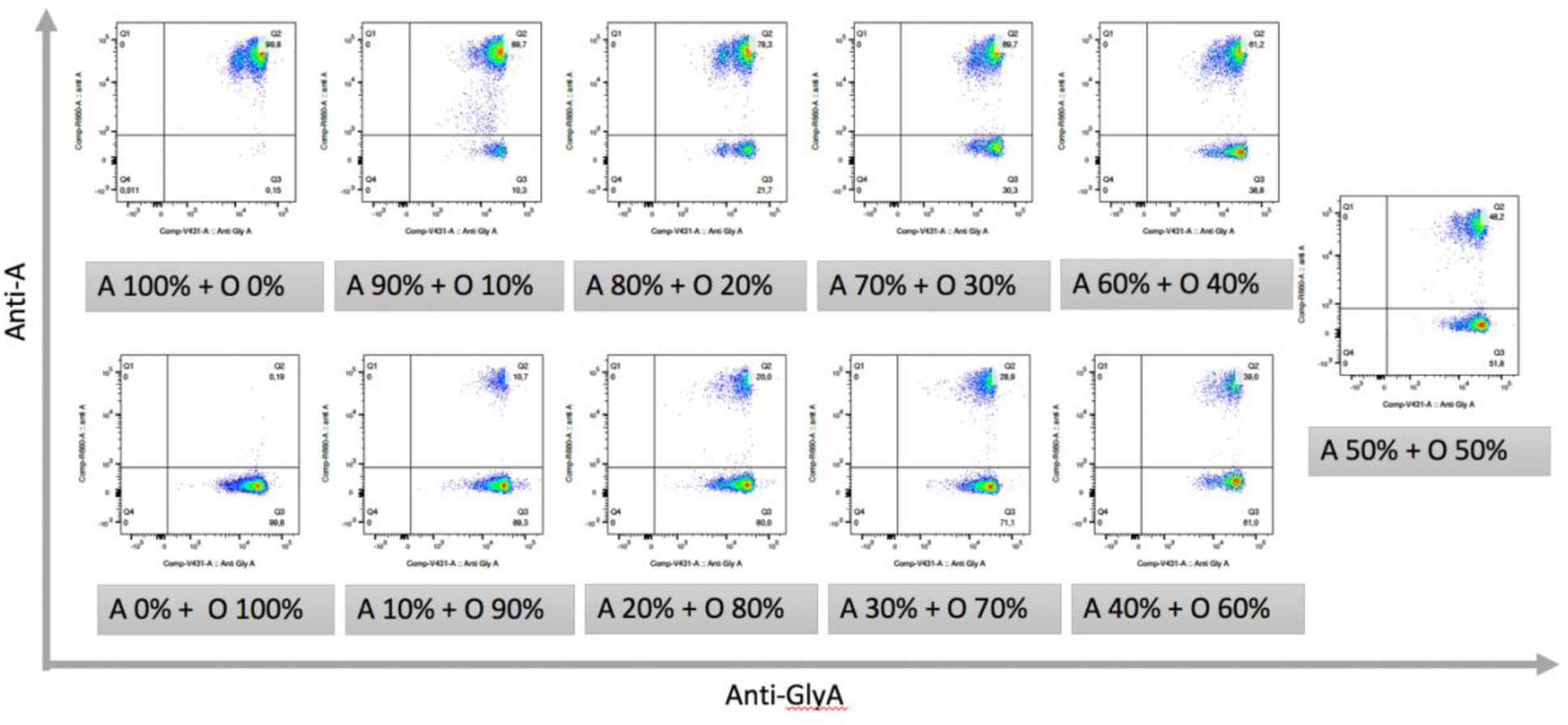
Flow cytometry data for A_1_/O – 2023-02-09. Data obtained through using the proposed FCM-based RBC chimerism assay for artificially mixed combination of blood groups A_1_/O. The experiment was carried out on 2023-02-09.

**Supplementary Figure 1.4:**
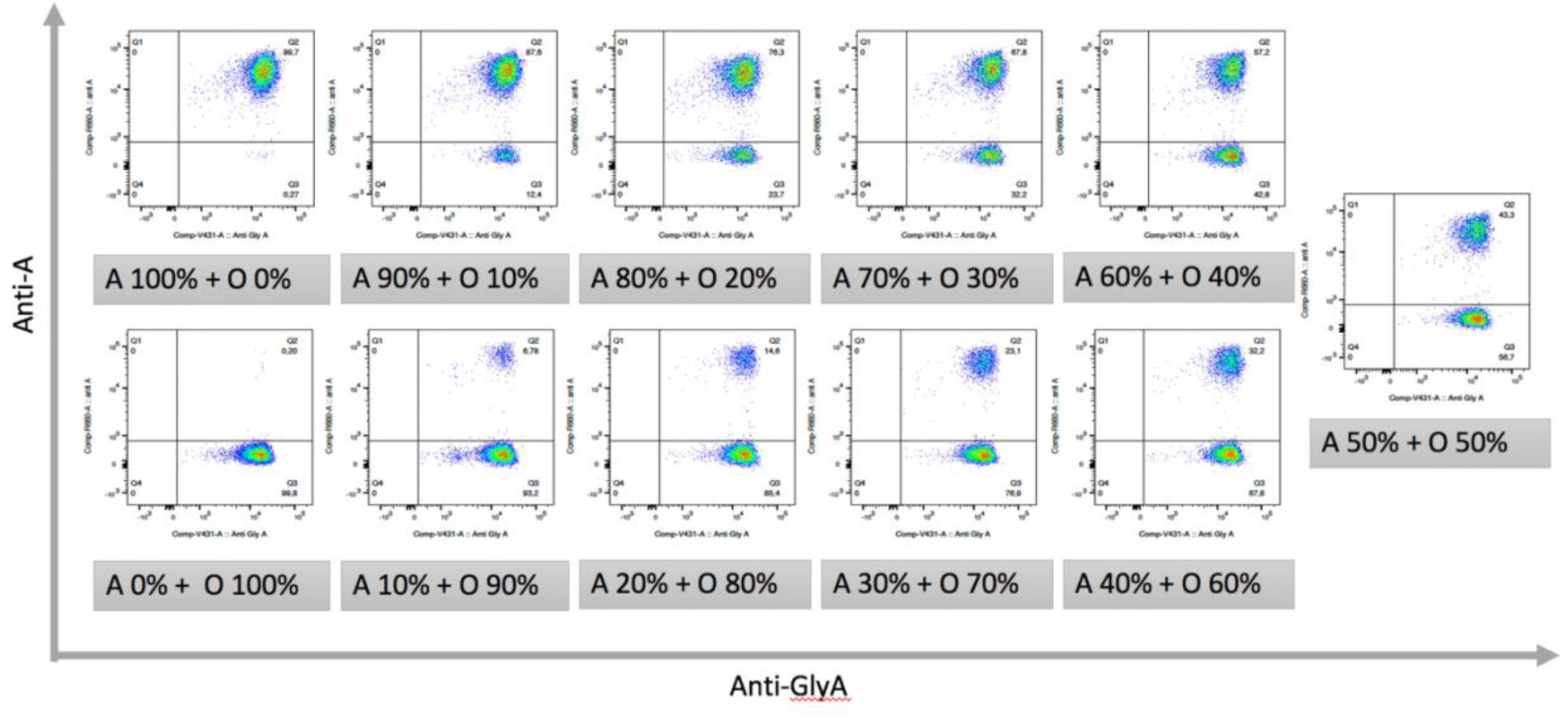
Flow cytometry data for A_1_/O – 2023-02-23. Data obtained through using the proposed FCM-based RBC chimerism assay for artificially mixed combination of blood groups A_1_/O. The experiment was carried out on 2023-02-23.

**Supplementary Figure 1.5:**
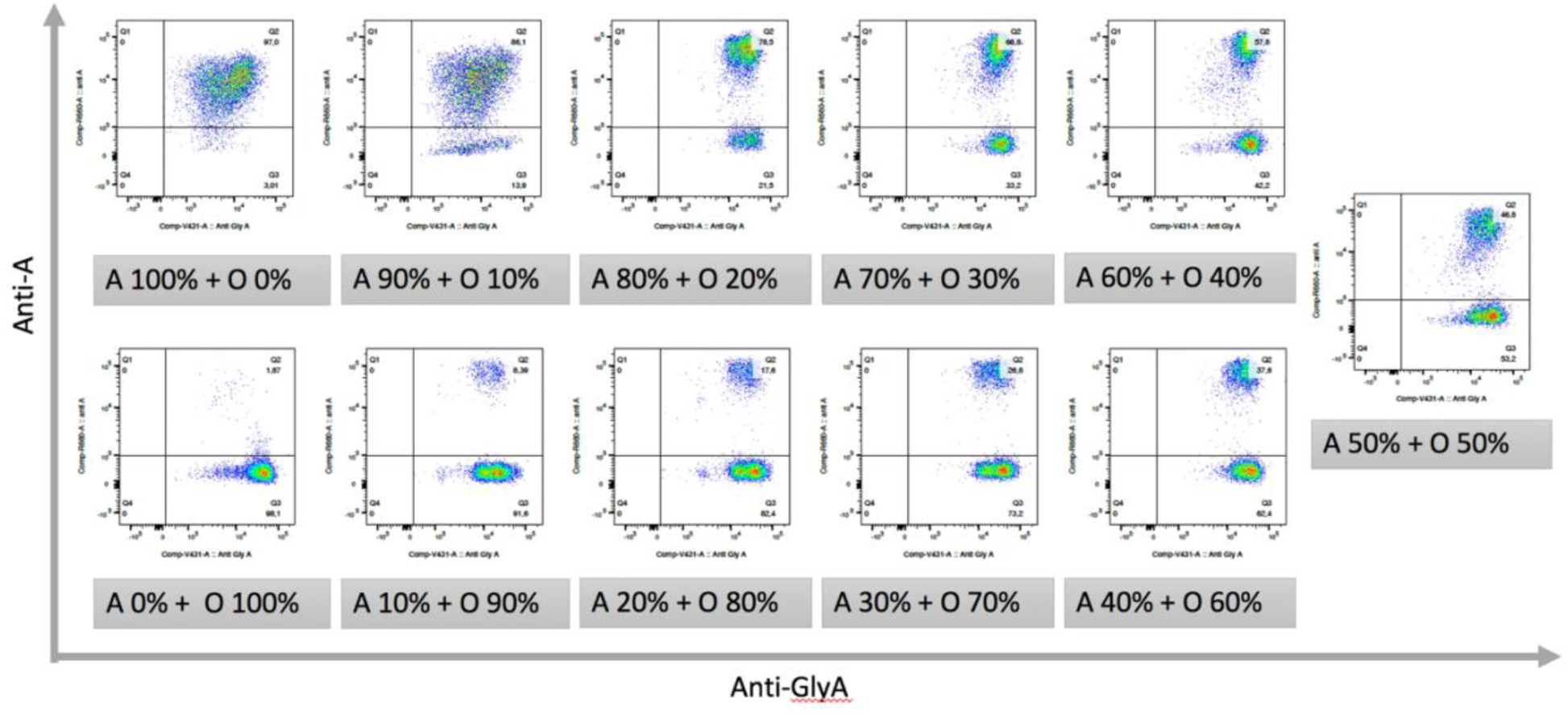
Flow cytometry data for A_1_/O – 2023-03-27. Data obtained through using the proposed FCM-based RBC chimerism assay for artificially mixed combination of blood groups A_1_/O. The experiment was carried out on 2023-03-27.

**Supplementary Figure 2.1:**
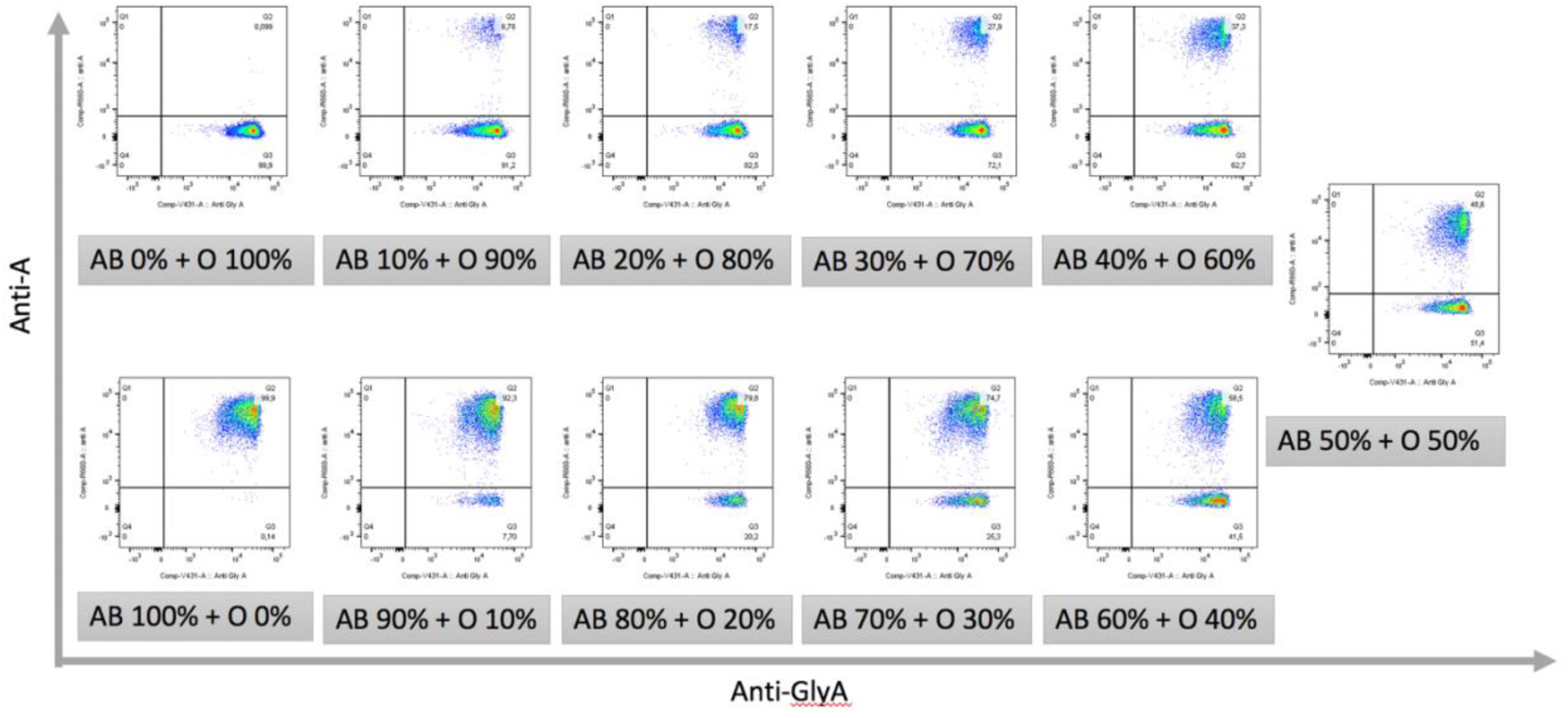
Flow cytometry data for A_1_B/O – 2023-01-19. Data obtained through using the proposed FCM-based RBC chimerism assay for artificially mixed combination of blood groups A_1_B/O. The experiment was carried out on 2023-01-19.

**Supplementary Figure 2.2:**
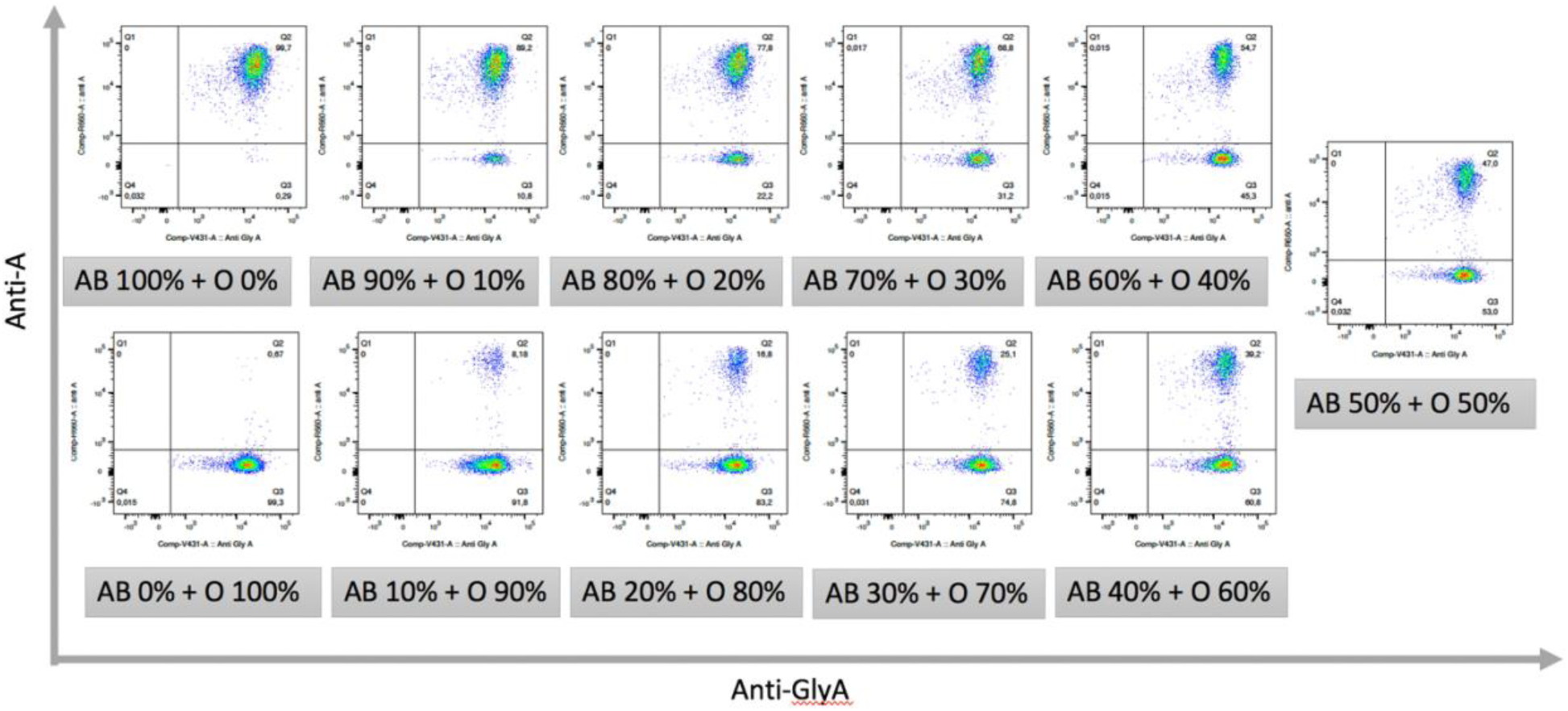
Flow cytometry data for A_1_B/O – 2023-02-03. Data obtained through using the proposed FCM-based RBC chimerism assay for artificially mixed combination of blood groups A_1_B/O. The experiment was carried out on 2023-02-03.

**Supplementary Figure 2.3:**
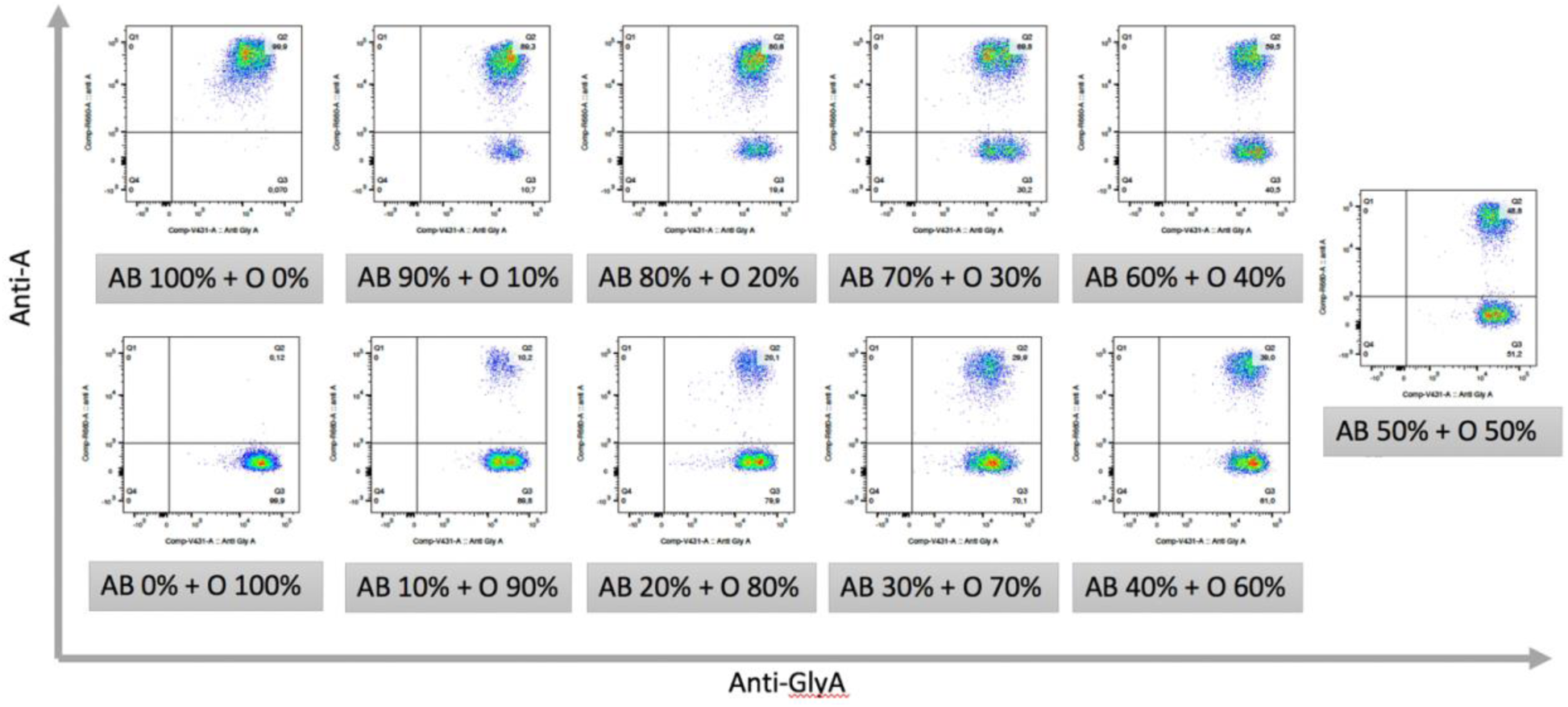
Flow cytometry data for A_1_B/O – 2023-03-16. Data obtained through using the proposed FCM-based RBC chimerism assay for artificially mixed combination of blood groups A_1_B/O. The experiment was carried out on 2023-03-16.

**Supplementary Figure 2.4:**
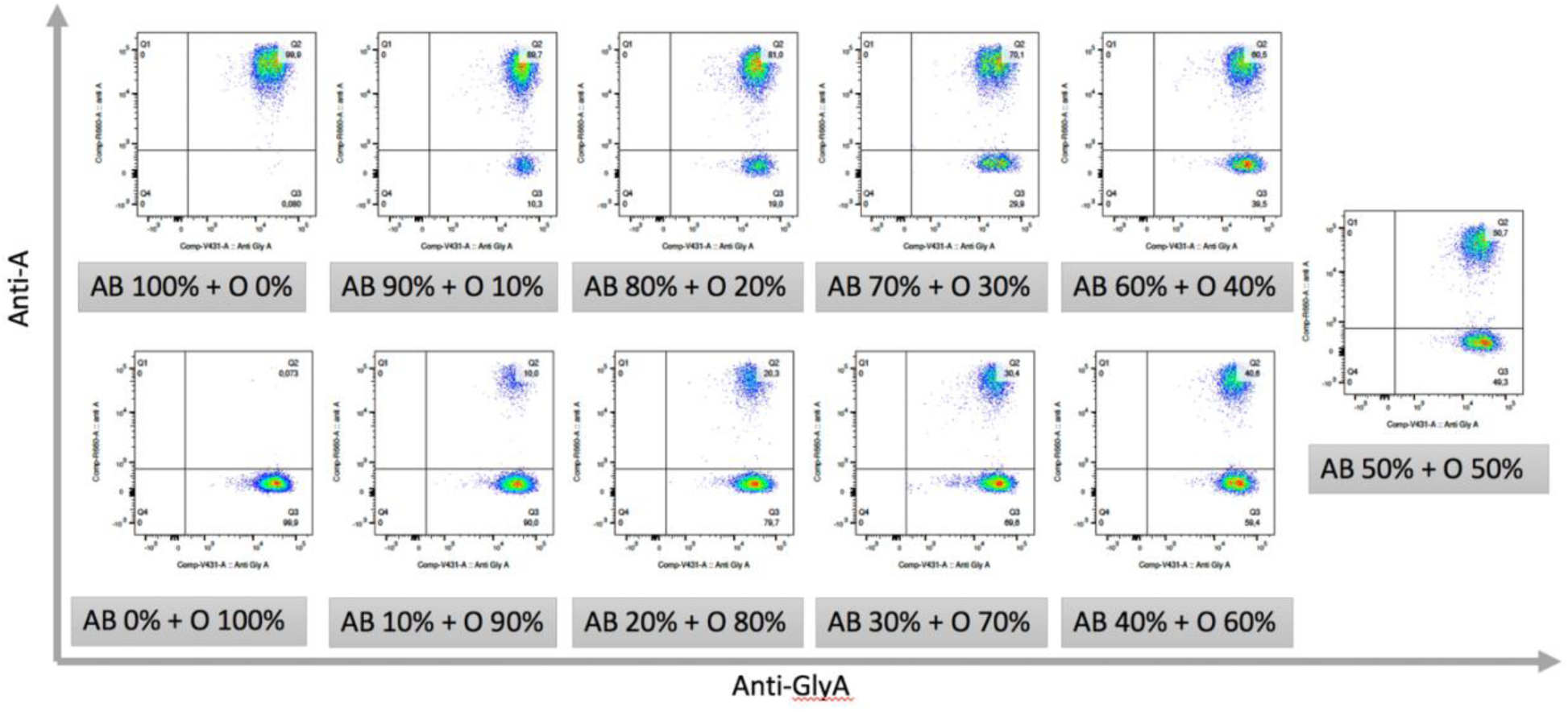
Flow cytometry data for A_1_B/O – 2023-03-16. Data obtained through using the proposed FCM-based RBC chimerism assay for artificially mixed combination of blood groups A_1_B/O. The experiment was carried out on 2023-03-16.

**Supplementary Figure 2.5:**
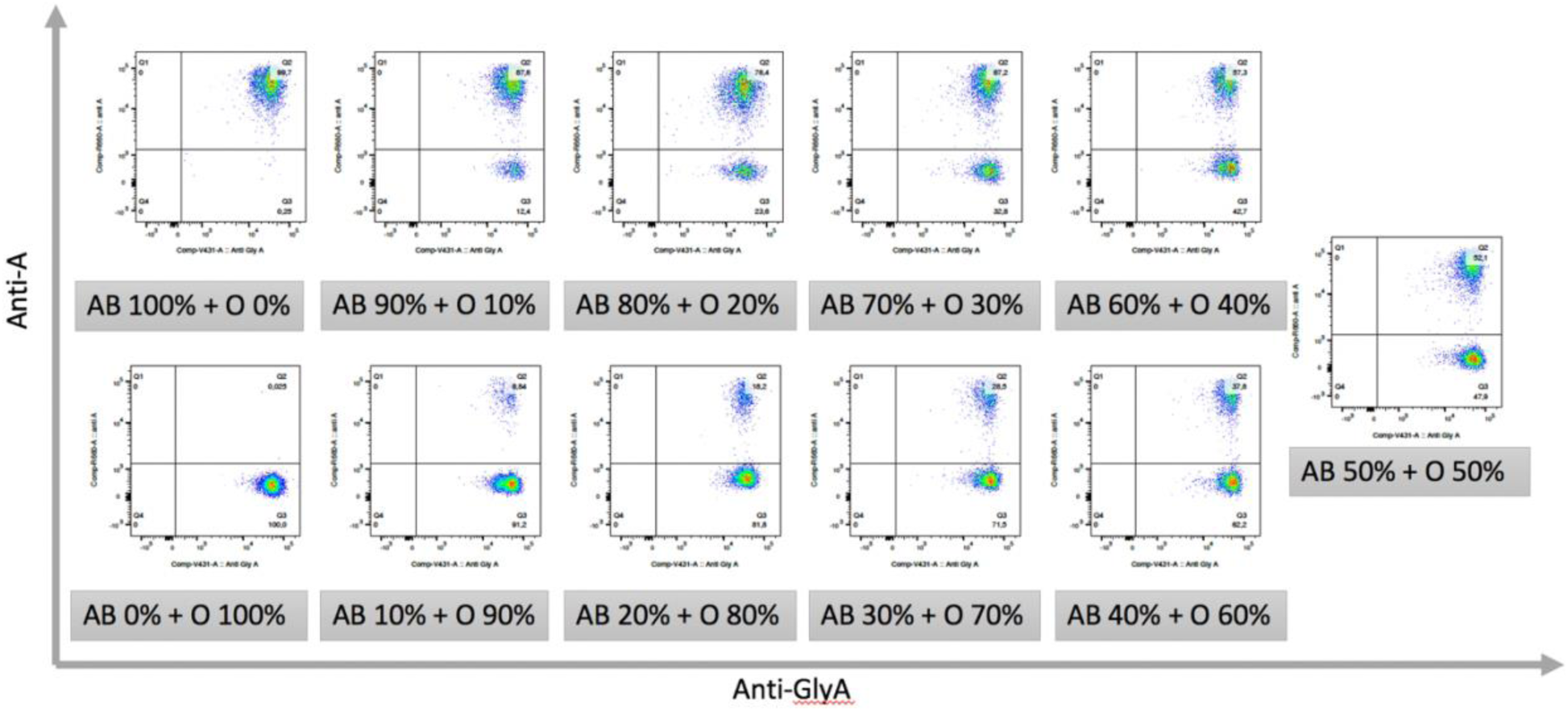
Flow cytometry data for A_1_B/O – 2023-03-27. Data obtained through using the proposed FCM-based RBC chimerism assay for artificially mixed combination of blood groups A_1_B/O. The experiment was carried out on 2023-03-27.

**Supplementary Figure 3.1:**
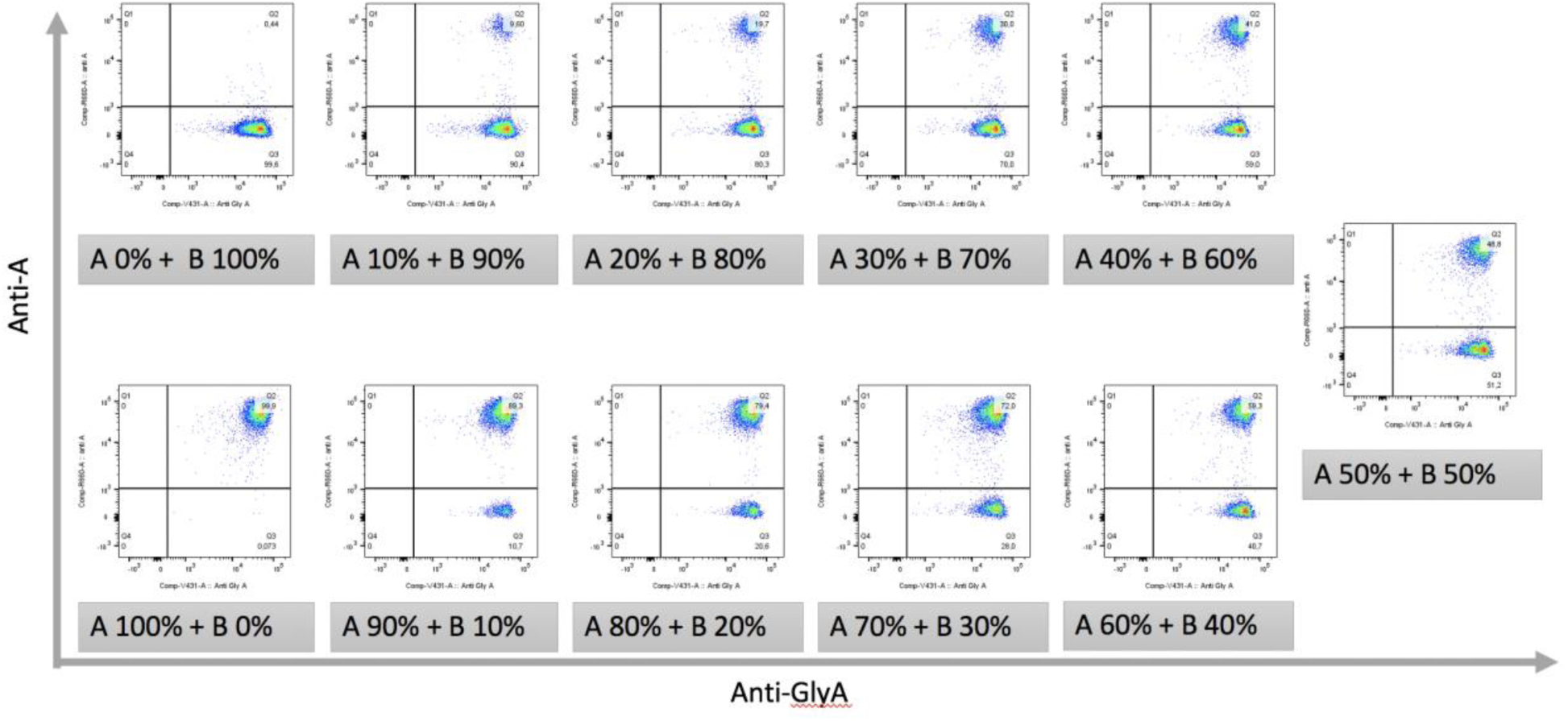
Flow cytometry data for A_1_/B – 2023-01-23. Data obtained through using the proposed FCM-based RBC chimerism assay for artificially mixed combination of blood groups A_1_/B. The experiment was carried out on 2023-01-23.

**Supplementary Figure 3.2:**
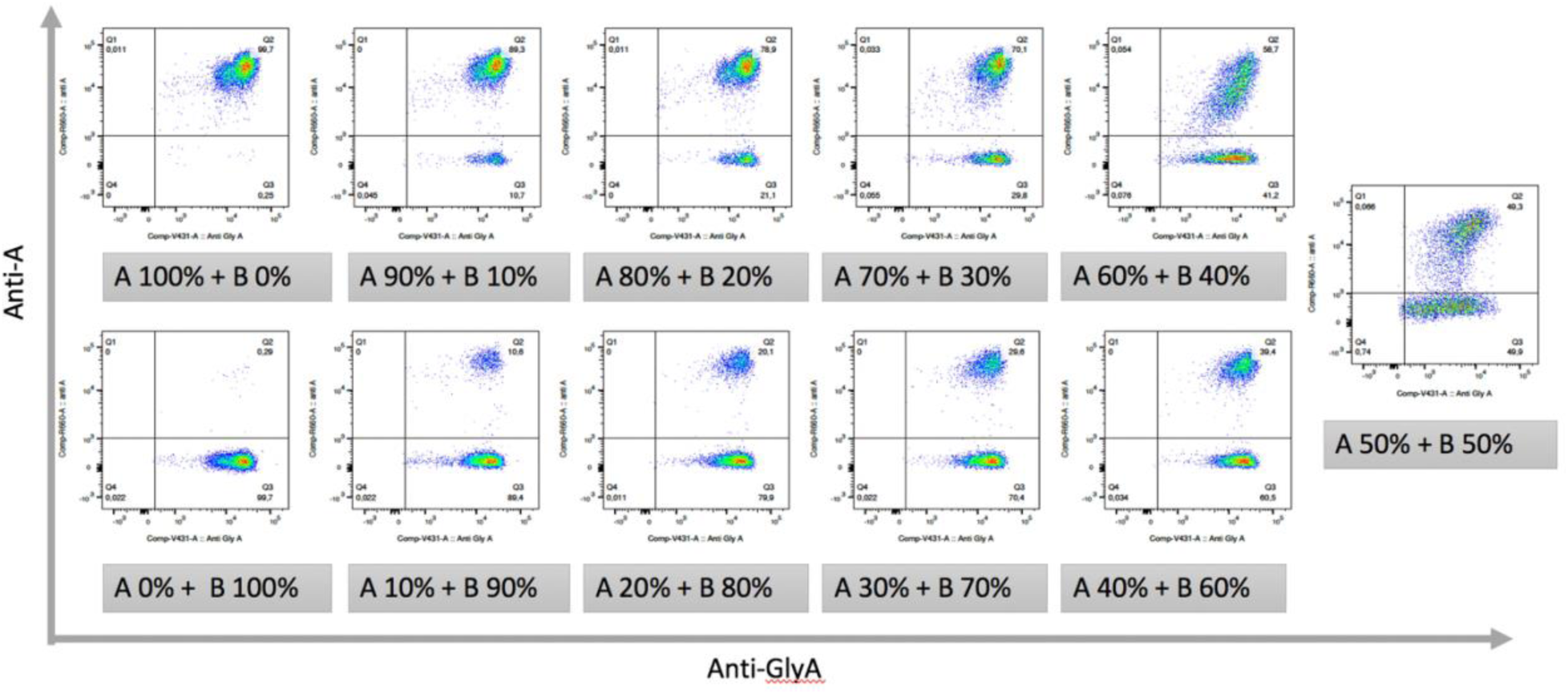
Flow cytometry data for A_1_/B – 2023-01-26. Data obtained through using the proposed FCM-based RBC chimerism assay for artificially mixed combination of blood groups A_1_/B. The experiment was carried out on 2023-01-26.

**Supplementary Figure 3.3:**
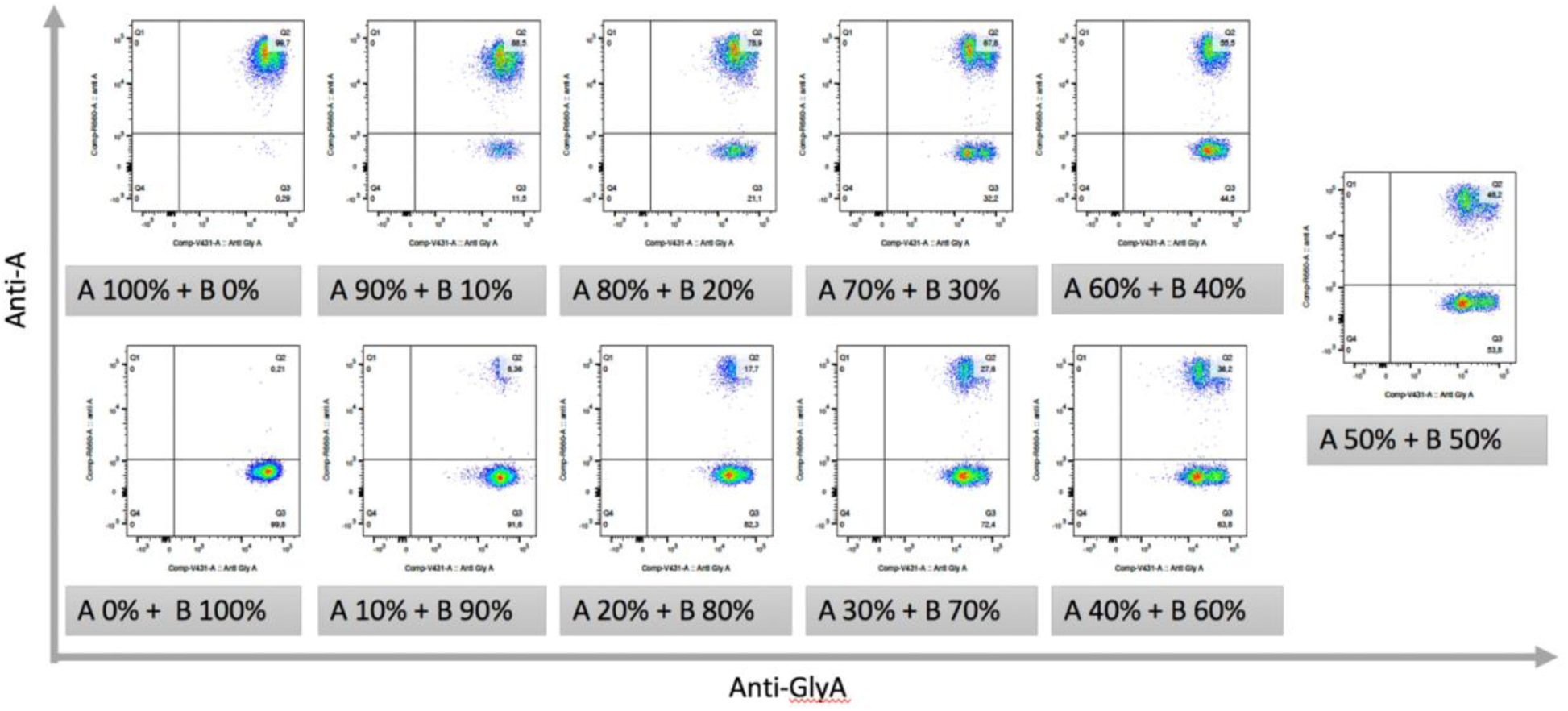
Flow cytometry data for A_1_/B – 2023-03-01. Data obtained through using the proposed FCM-based RBC chimerism assay for artificially mixed combination of blood groups A_1_/B. The experiment was carried out on 2023-03-01.

**Supplementary Figure 3.4:**
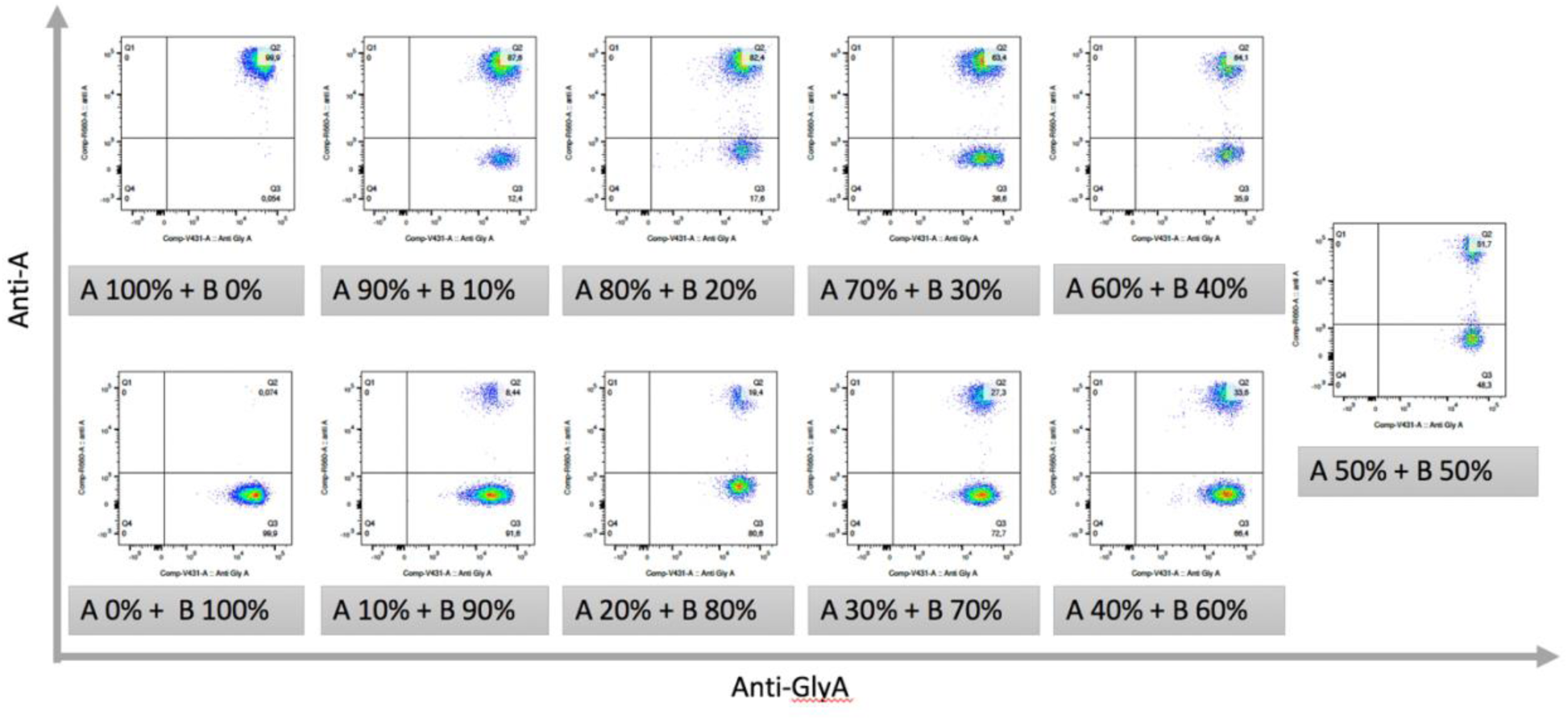
Flow cytometry data for A_1_/B – 2023-03-02. Data obtained through using the proposed FCM-based RBC chimerism assay for artificially mixed combination of blood groups A_1_/B. The experiment was carried out on 2023-03-02.

**Supplementary Figure 3.5:**
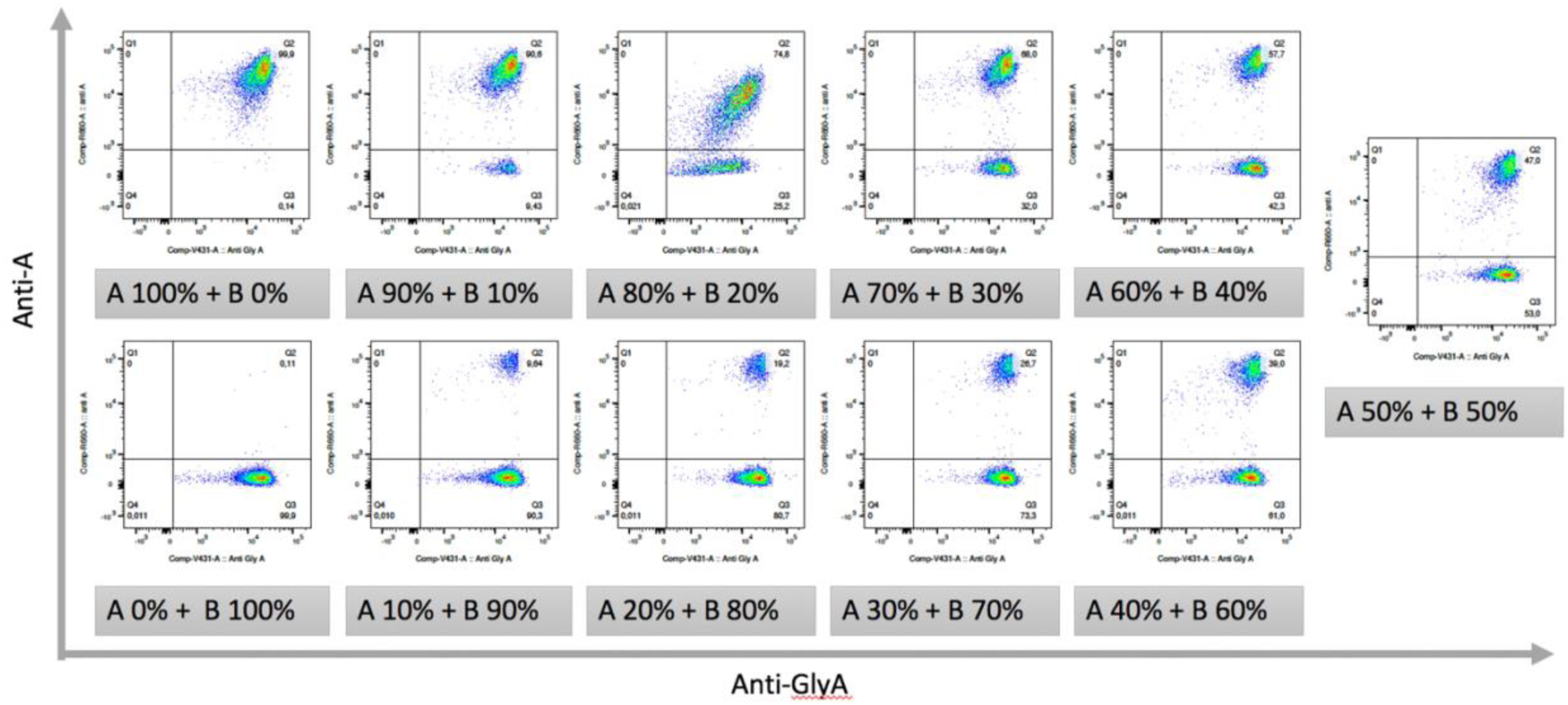
Flow cytometry data for A_1_/B – 2023-03-06. Data obtained through using the proposed FCM-based RBC chimerism assay for artificially mixed combination of blood groups A_1_/B. The experiment was carried out on 2023-03-06.

**Supplementary Figure 4.1:**
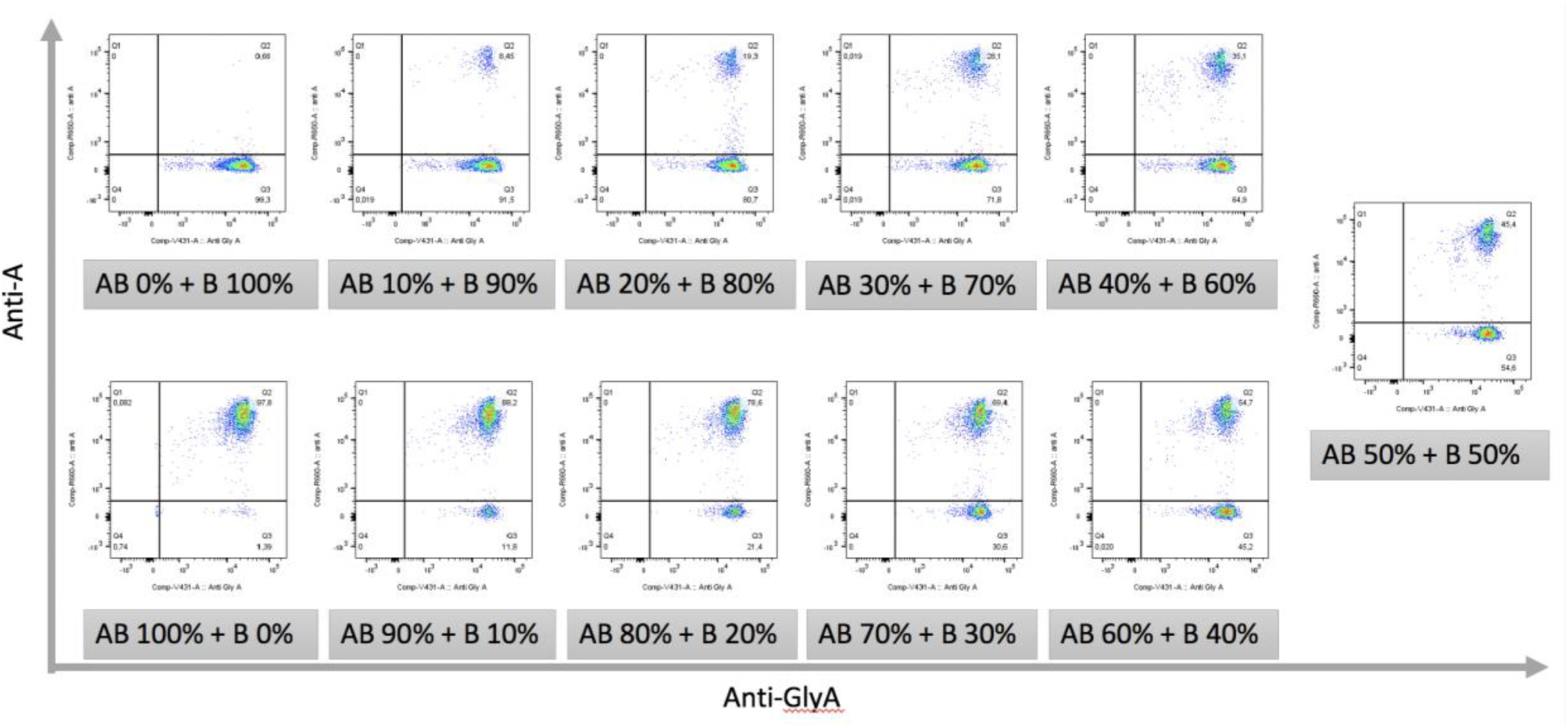
Flow cytometry data for A_1_B/B – 2023-01-20. Data obtained through using the proposed FCM-based RBC chimerism assay for artificially mixed combination of blood groups A_1_B/B. The experiment was carried out on 2023-01-20.

**Supplementary Figure 4.2:**
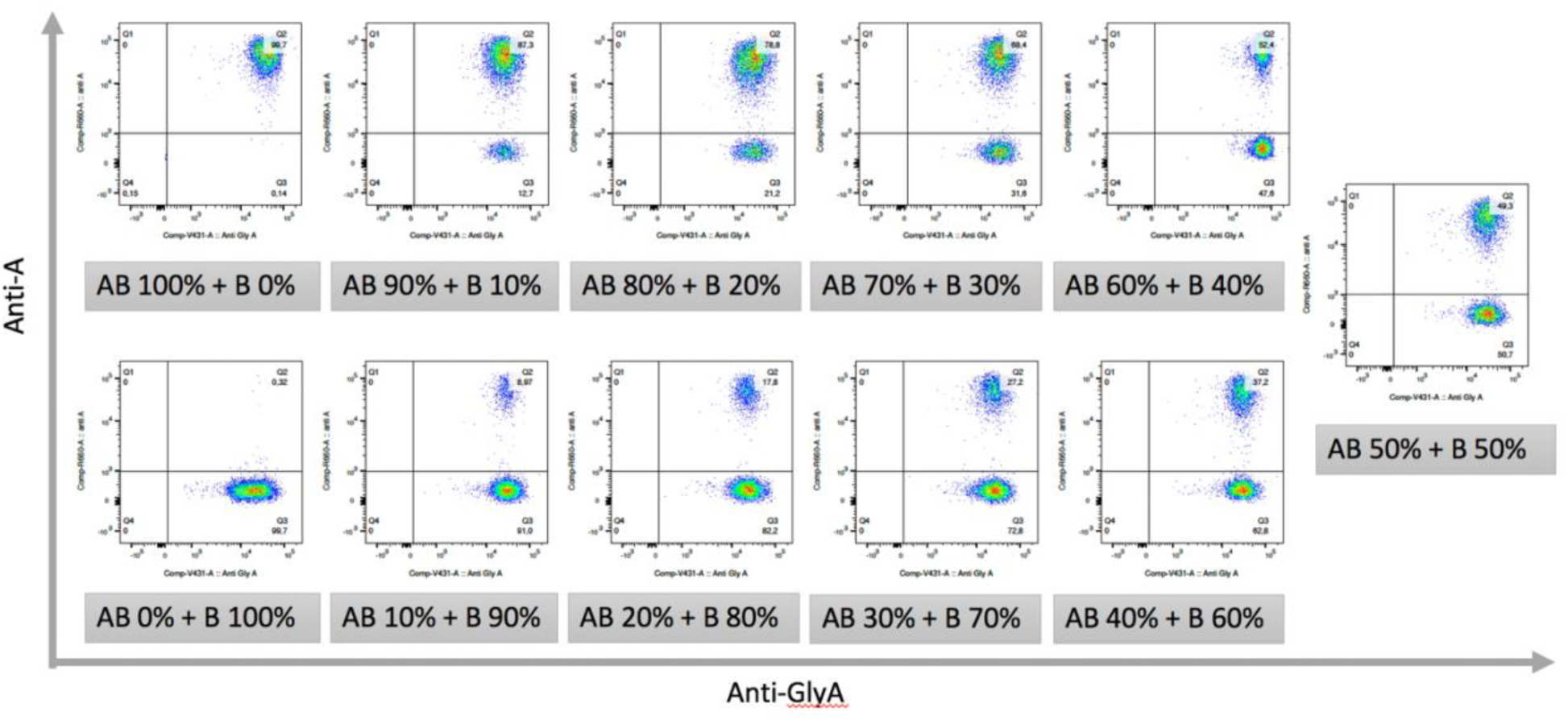
Flow cytometry data for A_1_B/B – 2023-03-15. Data obtained through using the proposed FCM-based RBC chimerism assay for artificially mixed combination of blood groups A_1_B/B. The experiment was carried out on 2023-03-15.

**Supplementary Figure 4.3:**
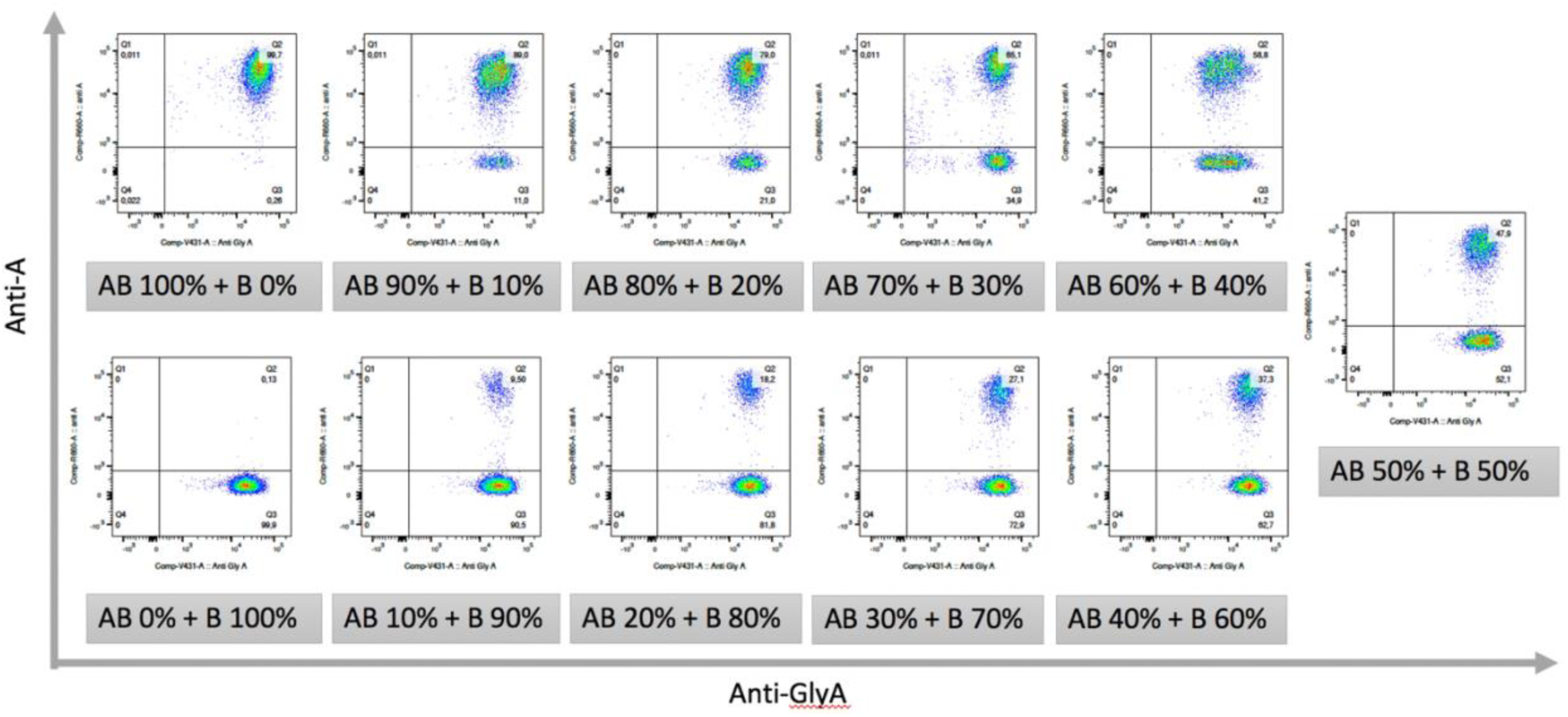
Flow cytometry data for A_1_B/B – 2023-03-15. Data obtained through using the proposed FCM-based RBC chimerism assay for artificially mixed combination of blood groups A_1_B/B. The experiment was carried out on 2023-03-15.

**Supplementary Figure 4.4:**
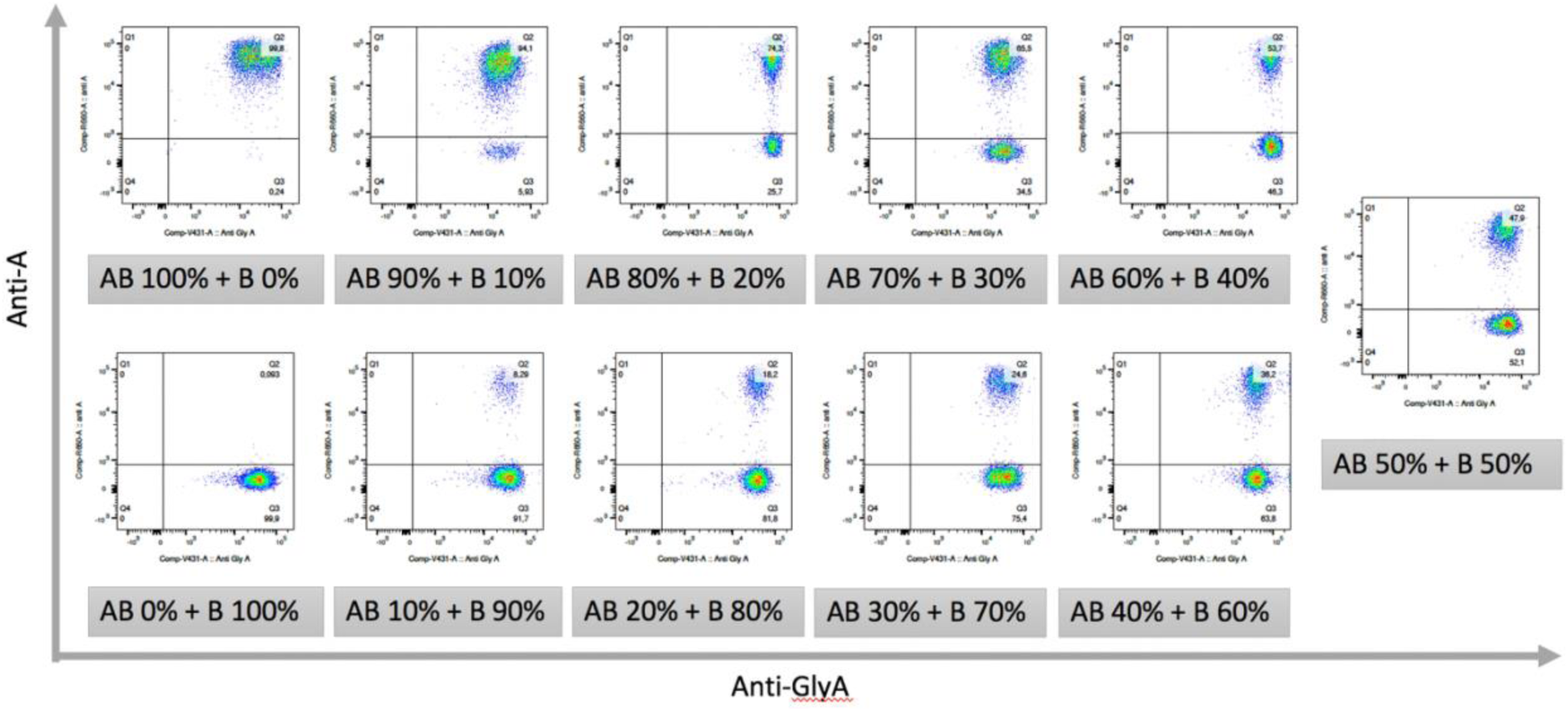
Flow cytometry data for A_1_B/B – 2023-03-15. Data obtained through using the proposed FCM-based RBC chimerism assay for artificially mixed combination of blood groups A_1_B/B. The experiment was carried out on 2023-03-15.

**Supplementary Figure 4.5:**
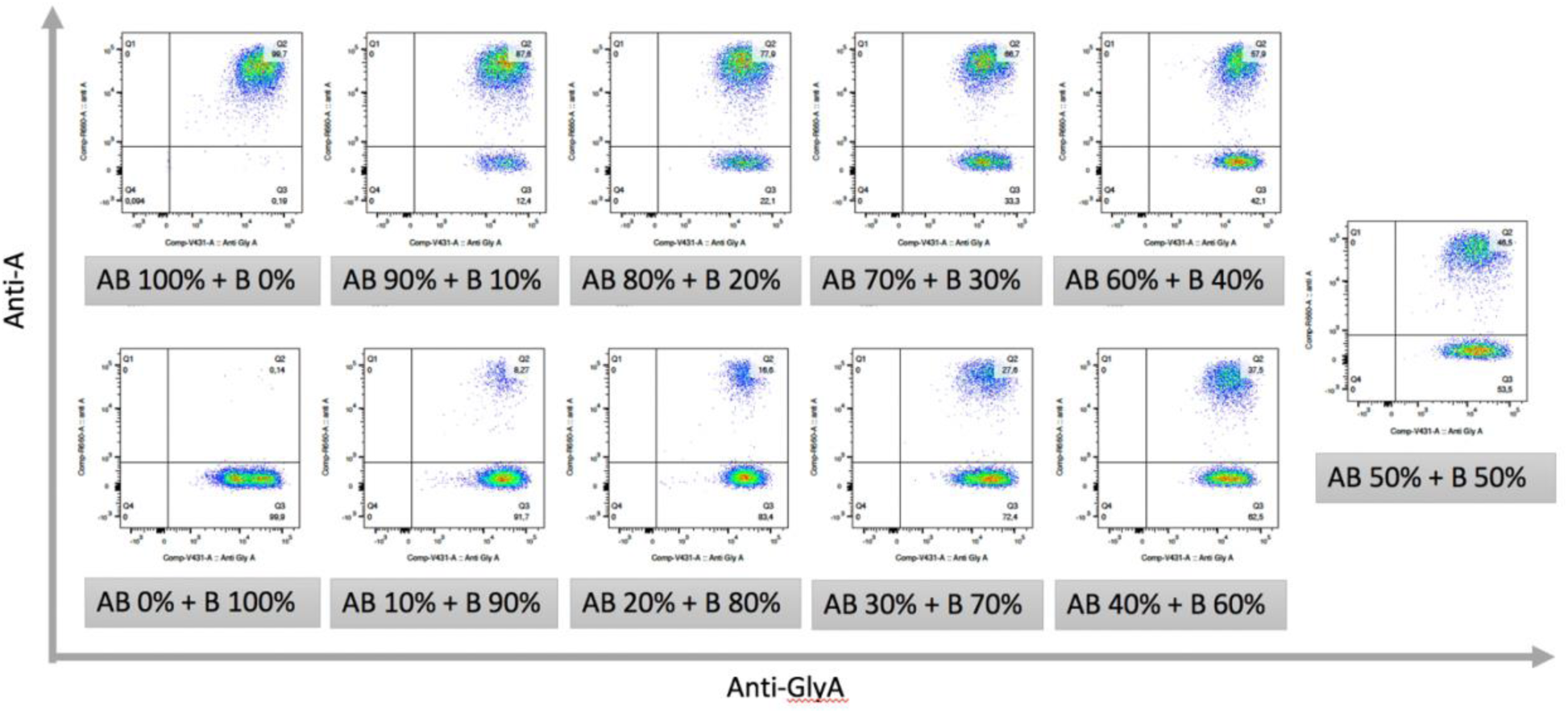
Flow cytometry data for A_1_B/B – 2023-03-16. Data obtained through using the proposed FCM-based RBC chimerism assay for artificially mixed combination of blood groups A_1_B/B. The experiment was carried out on 2023-03-16.

**Supplementary Figure 5.1:**
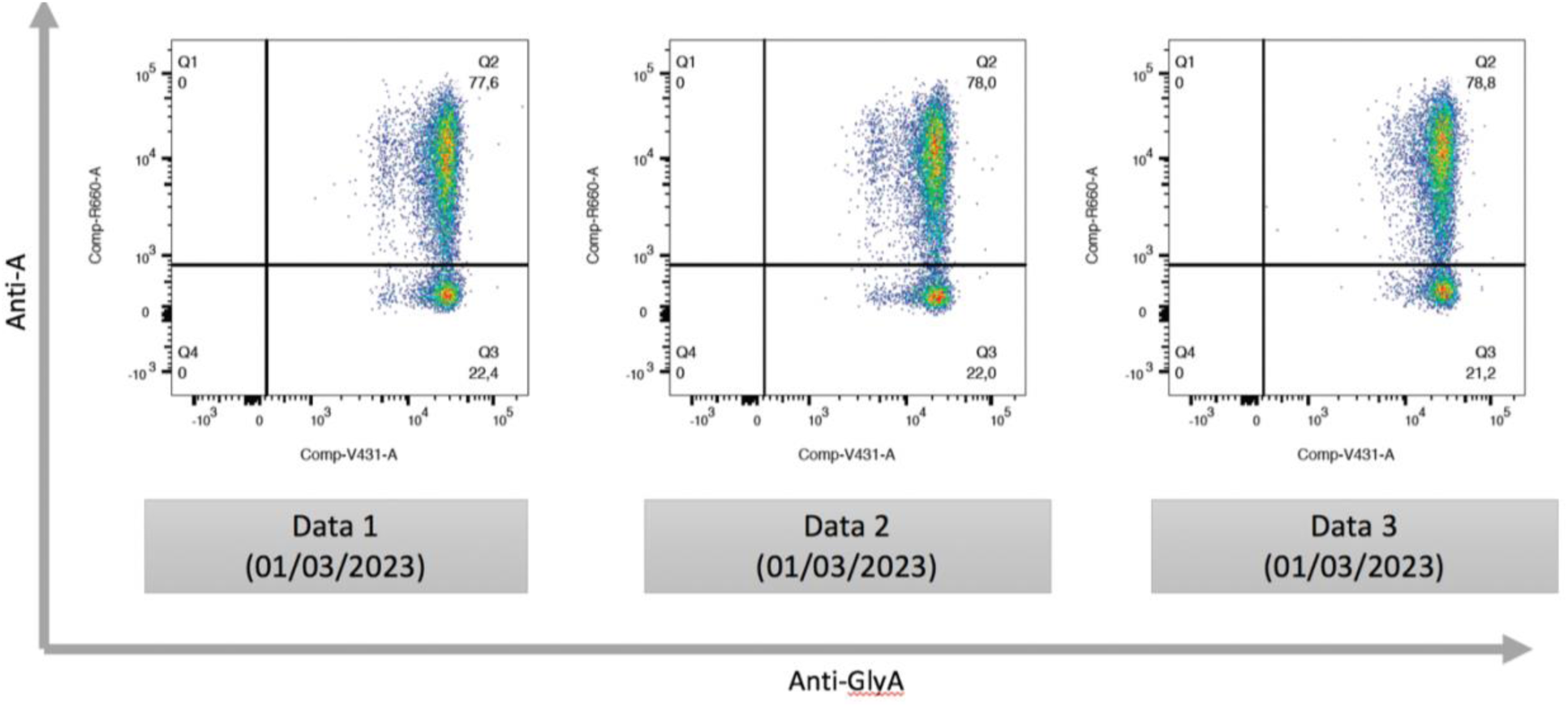
Flow cytometry data for patient sample with mixed chimerism of Recipient A_1_ and Donor O – 2023-03-01. A triplet experiment was done for the patient sample using the proposed FCM-based RBC chimerism assay. The data collected was shown in the figure. This experiment was carried out on 2023-03-01.

